# Decline in mitigation readiness facilitated second waves of *SARS-CoV-2*

**DOI:** 10.1101/2021.02.10.21251523

**Authors:** Kai Wirtz

## Abstract

Societal responses crucially shape the course of a pandemics but are difficult to predict. Mitigation dynamics is introduced here as an integral part of an epidemiological model, which is applied to the ongoing SARS-CoV-2 pandemic. Unperturbed simulations accurately reproduce diverse epidemic and social response trajectories from 2020 to 2021 reported from 11 European countries, Iran, and 8 US states. High regional variability in the severity and duration of the spring lockdown and in peak mortality rates of the first SARS-CoV-2 wave can be explained by differences in mitigation readiness *H* which is here mathematically defined as the value of human lives in relation to business-as-usual contact rates. *H* entails a suite of political, social, and psychological aspects of decision making. The simulations also suggest that a subsequent decrease in *H* much intensified the second wave and slowed down its decay. With less effective lockdowns, vaccination became the primary mitigation strategy in 2021. Retardation of vaccination relative to a 3-month scheme is projected to provoke an average toll of 1.5 deaths per million and delayed day. This toll particularly rises in regions with high numbers of old and still susceptible people, which is relevant for revising current policies of vaccine distribution.

---

Societies struck by the severe acute respiratory syndrome coronavirus-2 *(SARS-CoV-2)* pandemics in early 2020, mostly in Western industrialized countries, managed to reduce infection rates through non-pharmaceutical mitigation such as social distancing [*1,2*]. After these societies started to lift lockdowns in May 2020 [*3,4*], some reached very low case numbers, while others faced continuously high mortality caused by the coronavirus disease 2019 (COVID-19). Later in autumn and winter 2020/21, all these regions were hit by a second wave. Despite the experiences gained during the first lockdown [*5*], finding appropriate mitigation measures remained a difficult task [*6,3*]. The situation was further complicated in early 2021 by *SARS-CoV-2* spike mutations [*7*] and by the arrival of vaccines with uncertain distribution scheduling and public acceptance [*8*].

The lack of reliable mid-term future scenarios guiding the defense against *SARS-CoV-2* [*9,10*] stimulated the development of predictive tools. However, societal mitigation as a major control of the spreading dynamics was largely absent in classical epidemiological models. Simulation studies hence increasingly incorporated human agency [*11,12,13,14*], with diverse modeling approaches comprising rule-based, fitted, extrapolated, or pre-defined scenario settings [*15,10,16,17,18,19,20*]. Yet, no model could reproduce the observed re-adjustments in social distancing measures [*21,13*] across different countries in a forecast mode. Here, I seek to explain regional variability in viral spread dynamics and in societal responses by introducing an integrated societal epidemiological model. Numerical experiments covering the period 2020 to 2021 seek to provide insights and scenarios for supporting the ongoing battle against the pandemic and the attempts to reduce its devastating impacts on human lives and livelihood.

The mechanistic approach is built on a susceptible-infected-recovered (SIR) model, which resolves seven age groups, and is the first model that features (age-specific) contact rates as prognostic and adaptive variables. Adaptive changes in social mixing underlying the *SARS-CoV-2* transmission are here proposed to be driven by three pressures describing the benefits and costs of social distancing: (i) individual avoidance of own infection and mortality, (ii) social coherence in reducing overall infection levels, and (iii) costs of social distancing (see derivation in Methods). Changes in contact rates are then determined by the balance between the associated shift in COVID-19 mortality *M* – or the benefit of less mixing– and in the multifaceted socio-economic consequences *C* – or the costs of less mixing. Induced transmission shifts minimize integral social and mortality costs (*M* + *C*): during a pandemic, optimal contact rates are much reduced compared to business-as-usual (BAU) social mixing but still remain non-zero (Fig. 1a). For integrating *M* and *C* in the same metric, this work introduces the social trait *H* that quantifies the relevance of avoiding deaths versus keeping BAU contact rates (thus denoting a “human value”). This trait describes a full suite of aspects and dimensions in societal decision-making: the priority of governments to safeguard the economy, their facilitation of partisan polarization [*22*], capacity of elites and people to extrapolate in time (see Sec. S3), presence of misinformation and scepticism vs. effective science communication [*23,14*], group (in-)coherence and (non-)conformity to norms [*23*], individualistic vs. community oriented norms [*23*], psychological resilience vs. fatigue [*23,24*], or other individual attitudes such as patience, altruism, trust in institutions [*25,23,14*]. These aspects can in addition be mutually dependent such as norm adherence of individuals being linked to socio-economic inequalities [*24*]. In total, the aspects determine the *mitigation readiness* of a society during a pandemics. Societies characterized by a low mitigation readiness *H* tolerate a higher death toll before restricting mixing and mobility compared to those with a higher *H*. At low *H*, elevated social costs *C* curtail social distancing to small deviations from the BAU baseline (Fig. 1b). The mitigation readiness *H* is the only adjustable parameter of the model to address regional differences in the response to *SARS-CoV-2* throughout the entire simulation period. It is treated differently within two model variants: either *H* is kept constant at a base value *H*_0_, or steadily declines after the first lockdown from *H*_0_ at the degradation rate *r*_*H*_.

**Figure 1:**
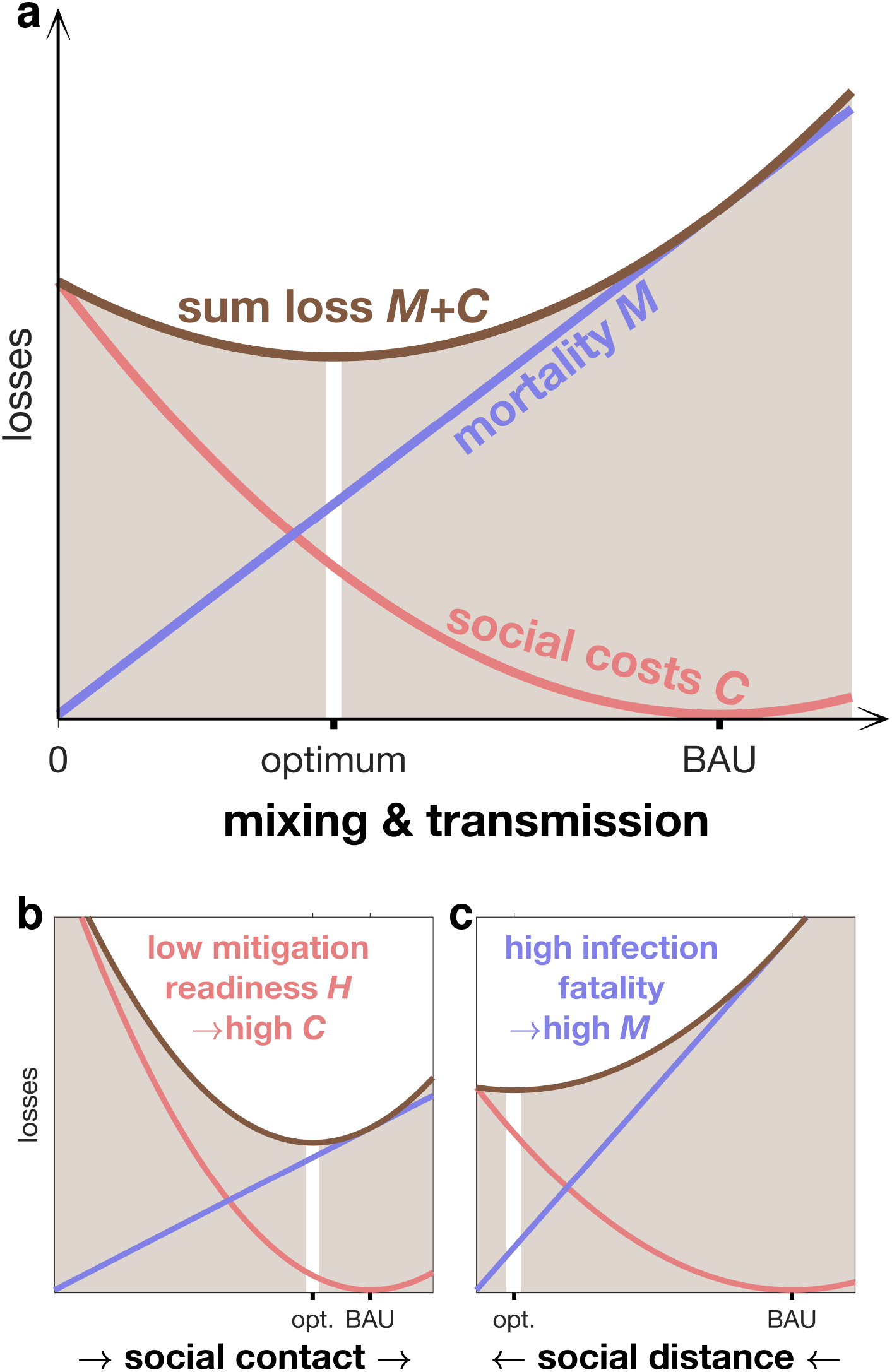
COVID-19 associated losses depending on contact rate. (a) Social costs *C* (red line) including, e.g., economic downturn, cultural loss, political instability, and psychological pressure, are inversely related to the value of social contacts vs. human lives (*H*). *C* is assumed to have a minimum at “business-as-usual” (BAU) contact rates and to increase non-linearly with growing distance from those BAU contact rates; mortality *M* (violet line) linearly increases with contact rate. The minimum of the sum loss *M* + *C* (brown line) is in the model approached by an adaptive adjustment in social mixing. (b) Younger societies will often feature a lower *H* due to lacking buffer mechanisms and lower fraction of people at risk compared to older societies. The resulting high social costs of social distancing keep the contact rate close to the BAU value. (c) In contrast, aged societies will have on average a higher infection fatality ratio and, concomitantly, mortality rate, which motivates stricter lockdowns.

In addition, fixed regional traits here describe differences in seasonality, BAU mixing patterns, and in age structure. Age affects epidemiological characteristics such as attack rates [*15,26,10*] and infection fatality ratio (IFR) which both are elevated in older age classes (Sec. S2). As a result, higher death tolls in aged populations will favor stronger social distancing (Fig. 1c).

At a given level of contact, viral transmission is assumed to decrease due to individual behavior and environmental factors. For example, moving everyday life outdoors or the wearing of face masks can effectively reduce exposure to viral infection (see Methods).

This study considers the COVID-19 associated mortality rate not only as part of the utility function but also as a major variable used for validation. Mortality data make a more reliable indicator for the infection state than the number of confirmed cases [*10,27,28*]. Selected by their high mortality rates in spring 2020, 20 regions were examined in this study, comprising 11 European countries, Iran, and 8 US states (see Tab. S1 and Methods).

## Model skill

Across the 20 regions, simulated COVID-19 associated death counts were consistent with the existing data (Fig. 2). Until Sep 2020, simulated mortality and data accurately match, and also the subsequent wave was reproduced by the model with only moderate deviations and time lags, including the occurrence of third waves for Louisiana, Georgia, and Iran. Fitting of the second and third waves can be further improved by calibrating three instead of one parameter (Fig. S1). The overall agreement is remarkable because mortality trajectories differed greatly among regions [*10,27*] and model runs represent true hindcasts: apart from a superimposed synchronous initiation of the first lockdown (see Methods) simulations were not corrected or tuned. This indicates a high predictive capability even in the mid to long term.

**Figure 2:**
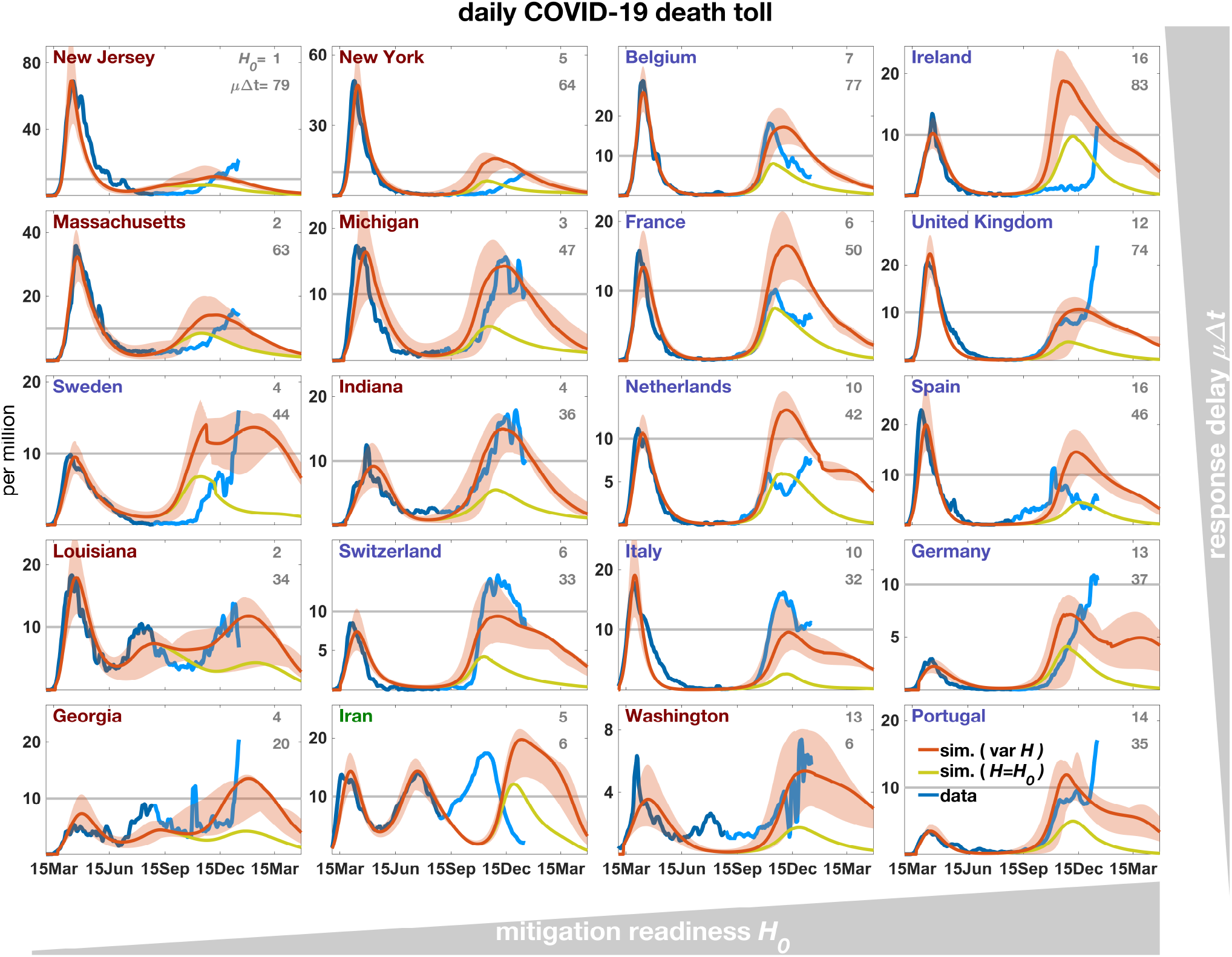
Daily mortality rate simulated either in a variant with constant *H*=*H*_0_ (olive line, *r*_*H*_ =0 in Eq.(4)) or one with decreasing *H* (*r*_*H*_ >0) after the first lockdown (red line). Uncertainty in model trajectories (shaded areas) arises from simulations with close-to-optimal *H*_0_ values as well as a range in external input (*γ*). From the reported and corrected mortality data (see Methods, blue line) only the first 180 entries were used for calibration of *H*_0_ (dark blue line), while the second half of the time-series is shown for comparison (light blue line). Note the different scaling of the y-axis as also visualized by the grey line at *M* =10^−5^d^−1^, which roughly corresponds to the mortality rate at starting capacity limitation of ICU hospitalization. European countries are labelled in blue, US states in red. The ordering of regions from left to right reflects increasing base *H*_0_ (defined in Eq.(5); grey numbers to the right top, relative to 10^4^), and from top to bottom the decreasing product of the initial spread rate 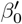 (Eq.(S6)) and the awareness Δ*t* (Eq.(S3)).

### Lockdown severity and mitigation readiness

The ratio between the reported intensity of social distancing and mortality during the first wave constrains regional values of the base mitigation readiness *H*_0_ (see Eq.(5) in Methods). High *H*_0_ were calibrated for many European countries that had faced a strong and enduring spring lockdown (Fig. 3) independent of their peak mortality rate (Fig. 2). To the contrary, inverse modeling attributed a relatively low *H*_0_ for most US states with their often milder lockdowns despite elevated mortality (Fig. 2, S2, Tab. S1). Values for US states, apart for Washington, lay in a narrow range (1.3–4.2 10^4^), which may point to a small variability of this aggregate social trait within countries. In regions with small *H*_0_ and lacking intense first lockdowns, mortality either decayed much slower compared to the average of all regions such as in Sweden, or a second wave built up already in summer 2020 such as in Louisiana (Fig. 2). The simulations well captured not only regional differences in lockdown severity, comprising a lockdown mobility above 50% of pre-pandemic levels (e.g., in Sweden or Georgia) or below 20% (e.g., UK or Italy), but also the different rates of recovery in mobility such as a fast return to BAU mobility in New Jersey versus a slower one in Washington (Fig. 3). The single calibration parameter *H*_0_ hence appeared to infer a realistic mutual interdependency of mobility and mortality patterns across regions so that mobility trajectories were overall in high quantitative agreement with the data.

**Figure 3:**
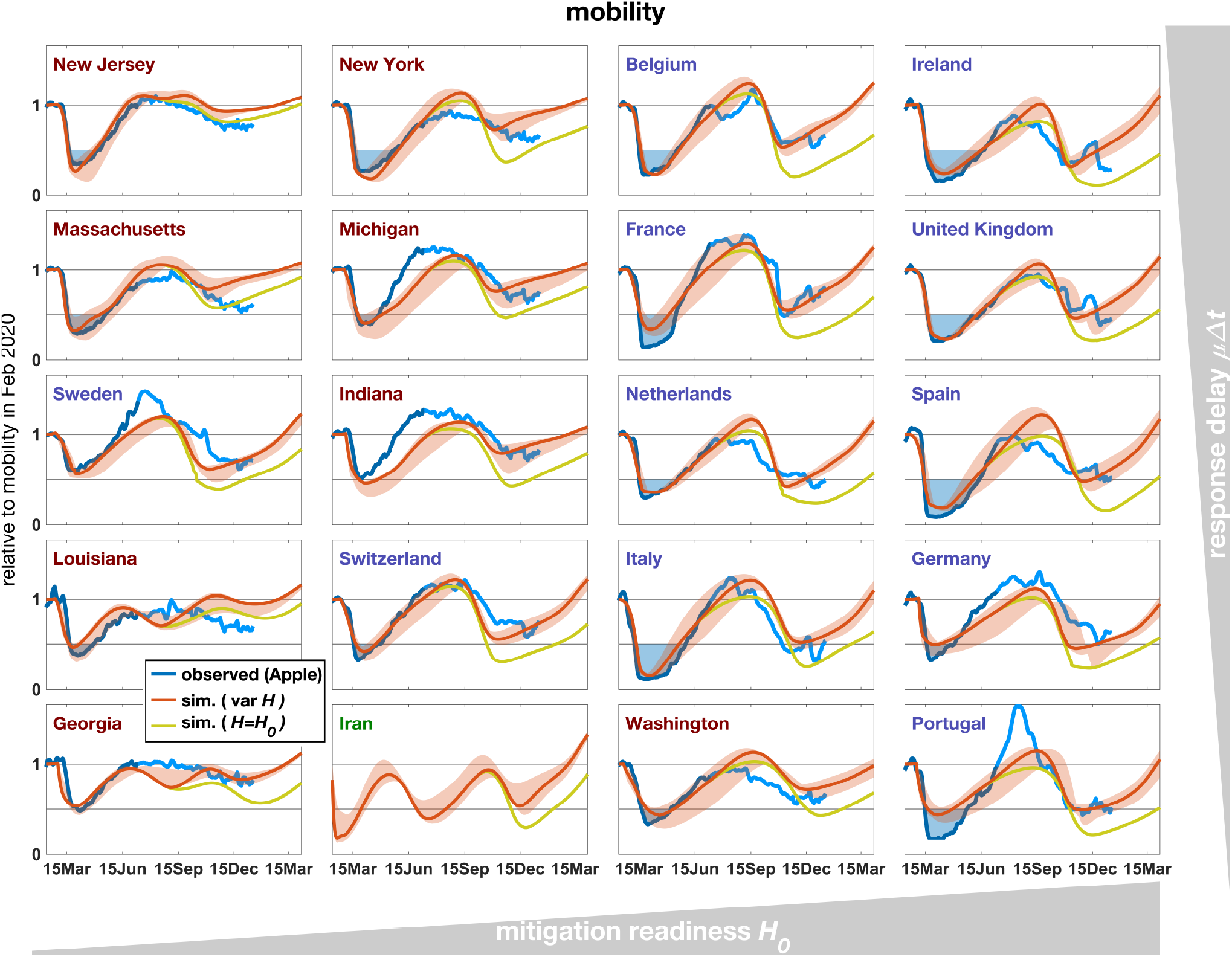
Mobility in 2020-2021 measured based on routing requests from mobile Apple devices (blue line), compared with the summed contact rate (Eq.(S8) in Sec. S7) in the reference simulation with constant *H*=*H*_0_ (olive line) and the simulation with decreasing *H* (red line, see Fig. 2). Severity of the spring lockdown is displayed as blue area below a mobility of 50% of the base level in Feb 2020.

### Decreasing readiness promoted the second wave

Contrary to first waves, second or third waves do not reach the actual death tolls if mitigation readiness stays constant in the model. Hindcasted peak mortality rates raising in autumn 2020 were on average by roughly a factor three lower than the reported ones. Only the model variant including a catchup mechanism generated by a steady post-lockdown decline in *H* (degradation rate *r*_*H*_ > 0, Fig. S2) enables a quantitative reproduction of peak death tolls, albeit in part with a temporal shift of up to 10 weeks such as for Ireland where data of late January (not shown) agree with the forecasted peak height (Fig. 2). Only for France and the Netherlands, the second wave seems to be better fitted by the first model variant, however at the cost of overestimating mobility in winter 2020/21 (Fig. 3). When extending the regional calibration to more parameters, COVID-19 mortality rates also of these countries were best reconstructed using non-zero degradation rates (Fig. S1). The model variant with *r*_*H*_ > 0 (*H < H*_0_) in general reproduces the strong social mixing during late 2020 in the data more accurately than the variant with *H*=*H*_0_ (Fig. 3). Better performance of the variant with *r*_*H*_ > 0 is also found for the third waves in Louisiana and Georgia (Fig. 2). These cases are particularly interesting to compare with an extensive US-wide study by the IHME forecasting team [*20*], which used a pre-defined scenario of mitigation measures. Peak mortalities of US states were either well predicted, or underestimated such as for Michigan, Indiana, and Massachusetts – or the two US states with a third wave (i.e. Louisiana and Georgia). For example, peak January mortality for Georgia reached 7 10^−6^d^−1^ in their reference run, in contrast to the approximately 20 10^−6^d^−1^ actually reported. The model presented here predicted 4 10^−6^d^−1^ when *H*=*H*_0_, but 14 or 20 10^−6^d^−1^ for *H < H*_0_ using the base or extended calibration, respectively (Fig. 2, S1). In the latter calibration, a late onset date (mid Nov) was used. This together with the initialization of the free IHME simulations at autumn 2020 point to a rather delayed and late decline of mitigation readiness in some regions (see also time lags for, e.g, Ireland, New York, or Sweden). The simple scheme proposed here (Fig. S2) thus requires refinements, which should also include mechanistic reasoning. Nonetheless, the overall enhanced model accuracy using a declining *H* can be interpreted as an indication for an actual relaxation to BAU normality, facilitated by the political, socio-economic, and psychological processes outlined above.

### Alternative pathways for industrialized countries

The moderate autumn/winter death toll in the model variant with constant *H*=*H*_0_ raises the question as to whether different mitigation strategies in the study regions could have led to practical extinction of the pathogen as realized by few Asian countries such as China [*29*]. The post-lockdown *H* was therefore shifted upwards in consecutive numerical experiments (and then kept constant). Increasing the mitigation readiness lowered the total post-lockdown death count; after raising *H* by about one order of magnitude, viral infection was eradicated across regions (Fig. S3, S4).

It can be doubted that Western societies would have tolerated deeper and longer cuts into individual rights of privacy and movement or into economic operations at nearly invisible infection density in summer-autumn 2020. However, magnitudes of the upwards shifts in *H* required for a full termination of the epidemics well correspond to the magnitudes of (dynamic) downward shifts reconstructed for the same period (Fig. S2). Hence, the necessary changes towards elevated mitigation readiness would not have been more radical *per se*, but directed towards the opposite direction compared to the actual decline in readiness.

Limited ability to fully prevent subsequent epidemic waves is implicitly hardwired in the model through the optimality assumption Eq.(7) targeting the least costly adjustment to the threat. While this approach is capable of “flattening the curve”, there may be more sustainable strategies aiming at total eradication of the pathogen [*29*]. A thorough “zero Covid” mitigation strategy is here induced by a huge value of every single case (*H* > 10^6^, Fig. S4), not necessarily because of the appreciation of the individual life (morality) but because of the exponentially growing number of –avoided– cases (see “expectation capacity” above).

Along these lines, in an otherwise non-preventive strategy also a full travel ban cannot much improve the situation. To the contrary, without imported cases, *γ*=0 in Eq.(1), simulated peak mortality rates of the second wave even increased in regions with low number of cases during summer (Fig. S5). This surprising phenomenon follows from the threat inherent to very low but non-zero case numbers at *γ*=0: When viral infection strikes from those very low levels, spreading rates can develop faster compared to the reference scenario (*γ* > 0). Yet, faster spreading rates are harder to defend against, which evokes higher peak mortality rates.

### Role of young people

Social distancing in the simulations similarly affected all age groups such that age distributions of cases were rather flat (Fig. S6), in qualitative agreement with first seroprevalence studies [*30,31*]. The decline of BAU contact rates from the younger to the elderly seems to be well compensated by the model setting of lower attack rates of the younger. As a result, young and medium aged cohorts can maintain finite contact rates during the lockdown, especially in low *H* regions such as the US (Fig. S7 and Fig. S6). Contagion within younger adults during summer 2020 fueled the epidemic rebound in all study regions (Fig. S8). The low IFR of young adults also explains why the ubiquitously higher case numbers of the second wave (Fig. S8) coincided with lower mortality in most of the regions [*32*].

The shift toward younger ages during summer is confirmed by US and German monitoring data [*33,34*], albeit there the cohort from age 15 to 30 (yr) appears most prominent whereas simulated infection levels were highest among adults older than 30 (Fig. S6). This discrepancy may indicate a lower conformity with mitigation measures within young cohorts than assumed by the optimal transmission regulation of the model, which is corroborated by studies revealing stronger non-conform attitudes among adolescents and young adults during the pandemics [*35,36,37,38*].

Higher behavioral exposure (*e*_*b*_) of young people together with their increasing dominance of the case distribution can induce a net shift in averaged *e*_*b*_ toward less protective behavior, even if the willingness to cooperate (e.g., by wearing face masks) stays invariant for each cohort as suggested by polls [*39*], redrawn in Fig. S9. This net exposure change was captured by the simulated relaxation of *e*_*b*_ (Fig. S9), although there is no explicit connection between age structure and behavioral changes in the model.

Changes in the frequency distribution inducing less defensive social traits (here represented by *e*_*b*_ and *H*) run contrary to selection for disease resistance as common in non-human populations. There, increasing dominance of more defended organisms reflects a correlation between infection and fitness. This correlation may be weak or absent in the case of the *SARS-CoV-2* pandemics affecting human populations, also because selective mortality of the elderly coincides with larger absence of multigenerational households, at least in Western countries (Sec. S7). Infected individuals younger than 50 not only experience a low risk of severe symptoms or fatality, but also exert little direct harm on their kin. Consequently, individuals with non-conform attitudes promoting exposure and susceptibility [*22*] lack the incentive to change these attitudes even after having been infected themselves – which actually is more likely than for conformists. This decoupling of variations in attitudes and their remote impacts may in part explain the reconstructed shift towards less defensive social traits in Western countries.

### Behavior and seasonality matter

During the course of the spring lockdowns, decreasing behavioral exposure *e*_*b*_ significantly helped to combat the first wave in the simulations (see also Fig. S10), a finding that underlines the relevance of using face masks [*40,41*]. The reduction of *e*_*b*_ occurred at different intensity among regions ranging from rather inert behavior (e.g., Iran, Georgia, or Sweden) to shifts by more than 50% (e.g., Ireland, Spain, or New York; Fig. S9). Readiness to improve behavioral protection appeared to increase under high peak mortality and/or high *H* value since both conditions cause intense (model) lockdowns that are here linked to behavioral shifts.

Even in regions displaying relatively inert behavioral adaptation, effective exposure *e* (= *e*_*b*_ *· e*_*E*_) markedly decreased in late spring 2020, which in the model follows from the transition to spread-reducing environmental conditions (*e*_*E*_). The decreases in *e*_*E*_ condense multiple bio-physical and behavioral processes driven by higher temperature and intensity of solar radiation such as effects on viral viability, or on placing activities from indoor to outdoor. Conversely, as also anticipated by virologists [*42,43*], returning autumn/winter conditions much contributed to the arrival of the second wave (Fig. S10), also visible from the synchronized dynamics of *e* and *I* (Fig. S9 and S8). Seasonality effects are, against expectation, most evident for regions at relatively low latitude such as Louisiana and Iran, where the increase in *e*_*E*_ already started during summer (Fig. S9) due to supra-optimal temperatures and was furthermore accompanied by high values of behavioral exposure *e*_*b*_ as mentioned above (see also Fig. S10).

### Inequality of vaccination and mortality rates

Higher behavioral as well as environmental exposure together with softer social distancing in winter 2020/21 considerably slowed down the decay of the second wave in comparison to the first wave (Fig. 2). In this situation, Western societies turned to vaccination to become the primary mitigation strategy as vaccines were approved and available from Dec 2020 onwards. However, vaccine rollout in 2021 will likely be hindered by, e.g., limited vaccine production, inefficient logistics, purchasing conditions, and low acceptance among the public [*8,44*]. All these factors differ between the study regions, not to speak of the announced completion targets of 3 months for USA and UK vs. 9 months for member states of the European Community. This motivated a set of scenario runs where the length of the vaccination period and the acceptance ratio was varied (see Methods). As expected, simulated death toll in 2021 increases with extending the vaccination period, and also with decreasing acceptance ratio (Fig. 4). A delay by 6 months in average costs nearly four times more lives, which is equivalent to 1.5 deaths per million and delayed day. For an aging country such as Germany this number amounts to 2.1 corresponding to an extra absolute loss of 178 deaths per delayed day, with a maximal mortality difference in March 2021 (Fig. S1). For comparison, a 30% drop in acceptance/efficacy within a 9-month scheme in average exacerbates the death toll by 17%, from 403 to 470 per million. Regional differences in death tolls projected for 2021 were found to cover a factor of about 8 from the lowest (Iran, Belgium) to the highest values (Portugal) at a 3 month period and a factor of 4-8 (Spain or Iran vs. Portugal) at a 9 month period. These stark differences mainly correlate with the product of (1) the mortality rate at vaccination start and (2) the fraction of susceptible and old individuals (Fig. S1). The large inequalities in vaccination effects question the prevailing vaccine partition among countries or regions as it neglects the current infection state of the population or abundance of the elderly that are still susceptible to *SARS-CoV-2*.

**Figure 4:**
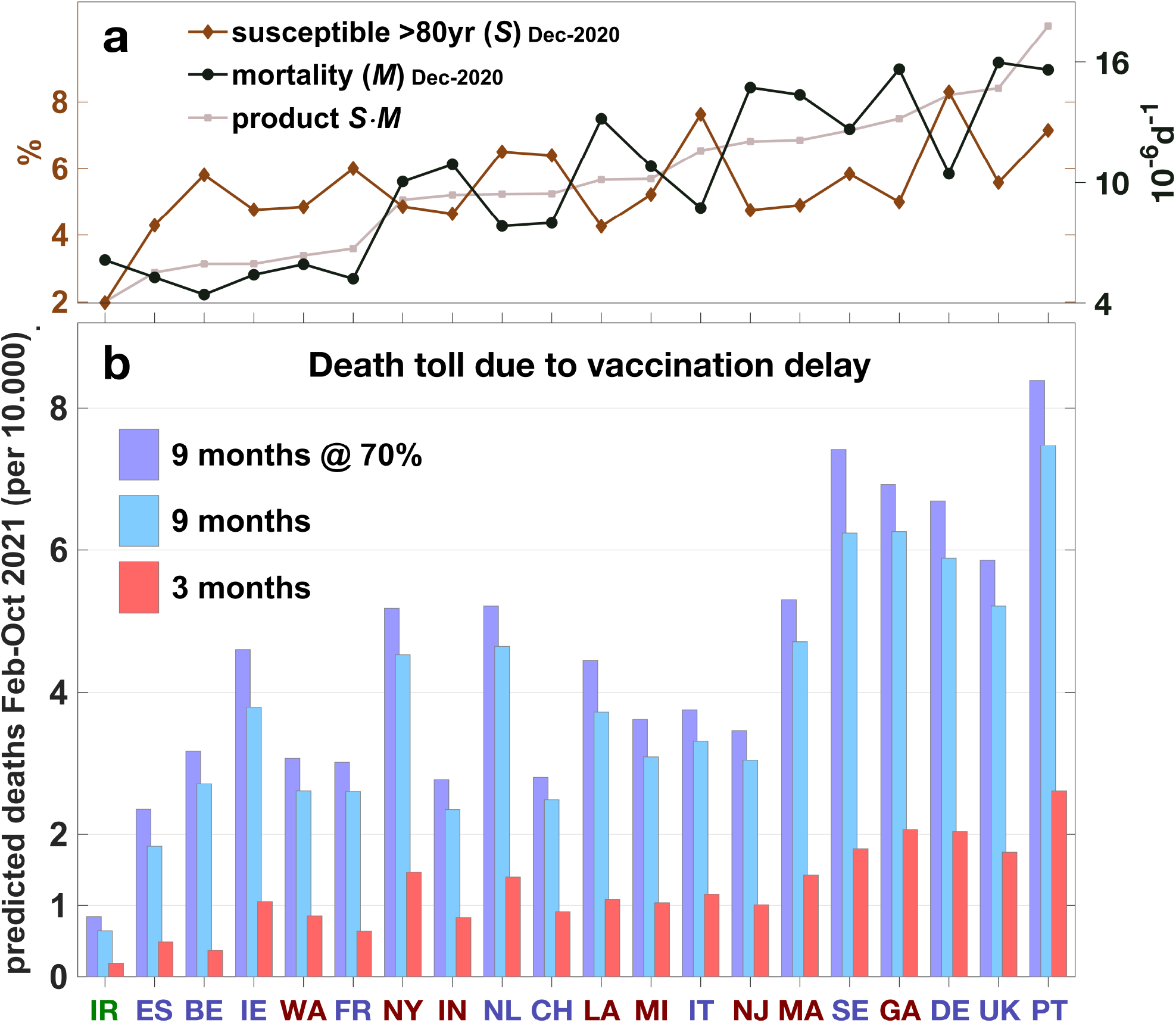
(a) Susceptible people in the oldest age group (*S*_7_, % of total population; brown line) and mortality rate (*M*, black line), both simulated for the vaccination start on Dec 25, 2020 (see also Fig. S1). The rescaled product of the two quantities (grey line) defines the ordering of regions from left to right. (b) Integrated projected death tolls from February to end of 2021 for three vaccination schemes characterized by vaccination period Δ*T*_vacc_ and vaccine efficacy: (1) Δ*T*_vacc_=3 months (100% efficacy, red bars), (2) Δ*T*_vacc_=9 months (100%, light blue), and (3) Δ*T*_vacc_=9 months (70%, purple).

### Short forecast horizon of state-of-the-art models

In all simulations, infection waves were halted by transmission reduction – or by vaccination – and not by depletion of susceptibles as forecasted by many state-of-the-art models. SIR models cannot seamlessly produce flat infection curves due to their mathematical structure combined with the lack of human agency, which in part explains why SIR models (alike statistical models) typically have a forecast horizon of only few weeks [*9,45*].

First attempts to extend epidemiological dynamics by macroeconomic factors [*46,26,47,25*] use a utility function similar to the approach presented here, and also distinguish between different types of agents such as “private individuals” (cf. here the selfish pressure) or the “social planner” (community pressure). However, economic models rely on equilibrium assumptions and on strictly quantifiable (monetary) units and, thus neglect potentially important non-economic aspects of societal decision-making such as learning under uncertainty, psychological fatigue, or political partisanism [*14,23,22*]. The negligence of sensible and dynamic control processes may be responsible for why the regular outcome of economic approaches remained within the herd immunization scenario of SIR models. In the presented model, the value of human lives (*H*) is defined in relation to an essential mitigation quantity during a pandemics which is social distancing, and not a monetary unit; furthermore, the results shown here suggest a high relevance for models to resolve societal responses dynamically.

Rather than social *dynamics* other recent approaches emphasize social *actions*: they are based on semi-heuristic rules of social distancing such as piecewise re-fitting of transmission [*48,19*], imposing pre-defined or rule-based shifts [*17,18,20*], relaxing transmission [*27*], and by relaxation cycles [*15,10*]. These approaches may be very supportive tools for short-term decision problems, but need to become more mechanistic with respect to their mitigation module, and also need to be validated at a monthly or longer time scale. More validation effort is also required for the model presented here, for example through applications to a broader range of societies, particularly those of non–Western countries, for testing model generality and suitability for supporting strategic planning. As for any model used for decision making, also this model has to be taken with caveats, which are briefly summarized in Sec. S10.

### Blueprint for adaptation problems

This study highlights social response and individual behavior and their possible deterioration as critical controls of the *SARS-CoV-2* pandemics. The unprecedented model skill over nearly one year across many regions may indicate (1) that the model captured governing principles of viral and social dynamics during a pandemic and (2) that societal responses display a high adaptive significance insofar minimizing both social costs and death tolls.

However, adaptive capacity is also an attribute of viruses. Mutations in *SARS-CoV-2* started to impact spread trajectories [*49,50,51,7*]. These mutational drifts in parameters of *SARS-CoV-2* virulence and incubation behavior can be resolved very analogue to the adaptive dynamics implemented in the social model (Eq.(7) in Methods). This extended framework would facilitate modeling studies on the evolutionary arm races between human societies and *SARS-CoV-2* or other viruses. The framework can further be used as a blueprint for related problems, such as Climate Change assessments, which share, e.g., the balancing of environmental pressures with costly adaptation and mitigation efforts, or the need for extrapolating aspects of the utility function into the future. During pandemics and Climate Change, human agency is not an external boundary setting but an integral part of the system dynamics.

## Material and methods

### The societal-epidemiological model

The epidemiological section of the model resembles a SIR model as it distinguishes between susceptible and recovered people and those infected by *SARS-CoV-2*. For seven age classes *i* = 1 … 7, it resolves the fraction of infected individuals *I*_*i*_ of age group *i* relative to the total population size. *I*_*i*_ increases when susceptible people in that age class (*S*_*i*_) contract the virus and decreases at specific recovery rate *r* (Tab. S2):

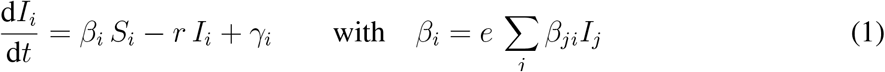

A global external input rate *γ* into a region (e.g., from travelers) is parametrized in Sec. S8. At simulation start, the fraction of susceptible individuals *S*_*i*_ equals the population fraction *ϕ*_*i*_ of the age cohort and thereafter declines due to infection and subsequent immunization or fatality,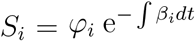. Group transmission rates *β*_*i*_ comprise variations in the effective exposure *e* = *e*_*b*_ *· e*_*E*_ by behavioral changes *e*_*b*_ and environmental factors *e*_*E*_ (see below and Sec. S9) and changes in contacts between age cohort *i* and all age groups. The specific transmission rate *β*_*ji*_ describes the probability *per individual* of potentially *contagious* encounter, and has to be distinguished from the contact rate *m*_*ij*_, which is the probability *per age group* of *physical* encounter,

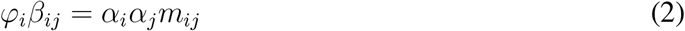

with specific attack rates *α*_*i*_ (Sec. S2). Infection described by Eq.(1) leads to a (lagged) mortality rate *M* caused by COVID-19 given by

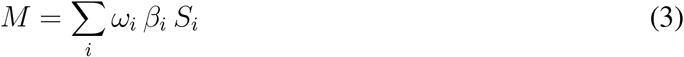

with age-specific IFR *ω*_*i*_ (Sec. S2).

Reductions in mixing and transmission by social distancing or other related restrictive measures induce a multi-facetted “social cost” (*C*) [*52*]. This quantity aggregates over various damages of social distancing on economic and psychological well-being, political stability, or cultural diversity [*1,16,53,52,54*]. Social cost *C* of mitigation is here assumed to rise with increasing social distance (denoted as SD), which sums over all differences of contact rates *m*_*ij*_ to their values *m*_*ij*,0_ before the epidemic, weighed by *m*_*ij*,0_ and sizes of interacting age classes.

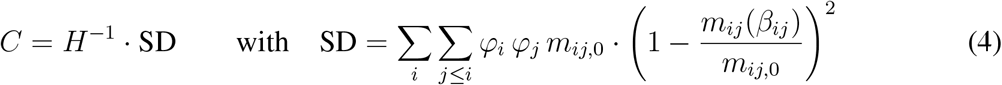

The quadratic dependency on contact rate ratios (being linearly related to *M*) resembles the relation between GDP loss and mortality at variable social distancing found by economic models [*26*]. It encompasses tolerance against small deviations but strong effects of downturning contacts to their minimum. The inverse proportionality coefficient, the mitigation readiness *H*, determines the height of mitigation costs perceived by a society in units of the mortality rate. Delayed and accumulating impacts on societal, economic, and psychological well-being [*55,53,56,54*] and consequential shifts in prioritization are here captured by a steady decrease of *H* at the “degradation” rate *r*_*H*_, activated on the day *t*_reset_ when net infection at low case numbers returns from a negative to a positive rate after the first lockdown,

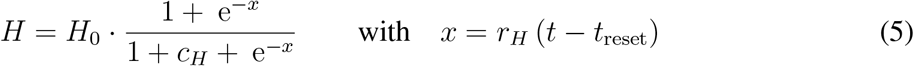

with reduction factor *c*_*H*_. The functional form derives from inverting (cf. *H*^−1^ in Eq.(4)) the logistic function that is a classical descriptor used in societal and economic theory [*57*]. A zero or non-zero degradation rate *r*_*H*_ distinguishes the two model variants used in this study.

*H* serves as a central linkage between the socio-economic part of the model and the epidemiological one or, more specifically, between the different meanings of the two loss functions *C* and *M*. This enables to define the total loss *L*, which as the utility function of the integrated model guides societal responses during the pandemic:

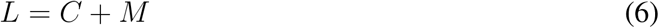

Avoidance of pathogenic transmission (by lowering *β*_*ji*_) and, as a consequence, reduced COVID-19-associated death toll has to be traded off with associated social costs. Societal transmission regulations are here suggested to be rational in terms of minimizing the combined loss *L*. The existence of the utility function *L*(*β*_*ij*_) allows to describe social regulations as adaptive dynamics of specific transmission rates *β*_*ij*_. Following the adaptive trait dynamics approach [*58*], once even applied to societal dynamics [*59*], this is formulated as an evolution equation for *β*_*ij*_ entailing a “responsiveness” *δ* times the marginal dependence of *L* on changes in *β*_*ij*_.

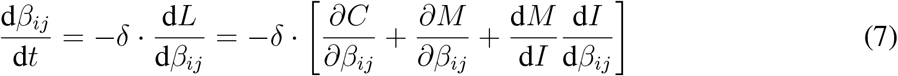

In a physical analogue, responsiveness *δ* describes the conductivity of how fast emerging threats induce new societal rules, and the bracketed derivative expression as a pressure acting on social traits, which is divided into three parts (see also Sec. S3): the first term in Eq.(7) can be directly calculated from Eq.(4) to be proportional to *β*_*ij*,0_ − *β*_*ij*_ and hence seeks to relax societal life to the pre-pandemic state. The second term in Eq.(7) quantifies the demand of life protection and simply follows from the mortality dependence on infection rates in Eqs.(1)–(3). This term is proportional to the IFR *ω*_*j*_ of the target age group, which strongly decreases in younger cohorts (Sec. S2). As a consequence, only interactions with and among senior groups would experience high reduction pressure; however, these contacts among or with the elderly cannot be shut down entirely (see Sec. S7), so that virulence among the young people can persistently contaminate the old ones. This side effect of isolated regulation in individual age-groups necessitates the extension of the adaptive dynamic framework by the third, “community-oriented” derivative term in Eq.(7) based on averaged target variables (*I* instead of *I*_*i*_). This term represents the responsibility of governments and the population as a whole, and requires sociality of young, non-risk groups (Sec. S3).

In addition to the adaptive shifts in contact rates, the model includes variations in the behavioral reduction of exposure *e*_*b*_. For example, wearing face masks or keeping sufficient interpersonal spatial distance up to self-isolation further lowers the infection risk at a given frequency of physical contact. The difficulty in formulating a reasonable cost function for behavior changes leads to a heuristic dynamics linked to social distancing (SD, defined in Eq.(4)): people are assumed to be more prone to adopt new behavioral rules at higher reductions in mobility and livelihood. This is expressed by a relaxation where *e*_*b*_ seeks to approach a target value *e*^*\**^ that decreases from its pre-pandemic value *e*^*\**^=1 with increasing SD

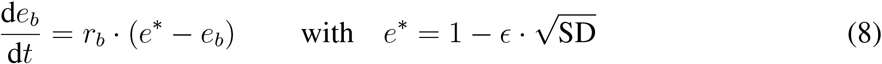

with specific adoption rate *r*_*b*_ and specific behavioral sensitivity *ϵ*. The square root dependency reverts the squaring in Eq.(4) in order to create sensitivity already to small variations in SD.

### Data integration and region selection

Fatality data were downloaded on Jan, 16, 2021 from the Johns Hopkins CSSE COVID-19 Dataset and smoothed by 7-day averaging. A regional correction factor was applied that averages the temporal means of the CSSE data and of the estimated excess deaths for US states [*60*] and for European countries [*61*]. Regions were selected if they had >700 death cases by April 25, 2020, and a relative mortality above the threshold *M*_crit_=7*·*10^−7^d^−1^ by Mar, 25, 2020. China was excluded owing to data irregularities and to its pioneering role in handling the epidemic. For Ireland, old cases from retirement homes reported on April 24 were re-distributed to the preceding time series. Iranian mortality data were multiplied with a higher and initially dynamic correction factor to comply with media reports [*62*]. Tab. S1 provides a full list of countries and states, correction factors, and demographic or regional characteristics.

For all study regions except for Iran, mobility has been reported from routing requests of Apple mobiles [*63*], which is taken as a measure for the intensity of social distancing [*64,65*]. For 7 of the selected European countries and USA at the country level, survey data on the willingness to wear face masks in the public [*39*] were used as a qualitative proxy to compare with simulated changes in behavioral exposure.

### Numerical experiments

This study is based on a systematic model calibration and three numerical experiments:

A. For each region, the model was run over 400 days from 21 days before the date when reported daily mortality matches *M*_crit_. Initial cases *I*_*i*_(0) were set proportional to (i) the regional age distribution *φ*_*i*_ and (ii) the critical onset mortality *M*_crit_. Initial transmissions *β*_*ij*_(0) = *β*_*ij*,0_ were derived from reported age–contact data and corrected using the slope of the mortality curve at the start of simulation (Sec. S6). The social trait *H* and the awareness factor Δ*t* (Sec. S3) were systematically varied in 800 simulations for each region. Epidemiological parameters were estimated from literature sources (see Sec. S2). The calibration of Δ*t* assured a rather synchronous lockdown timing of Western industrialized countries in mid-March [*66*]. The reported lockdown onset was anticipated by one week for Italy and Iran, and delayed by 5 days for all US states. Best fitting *H*_0_s were retrieved according to minimal root-mean-squared (RMS) deviation to mortality and mobility data while only the first 180 days of data were used. *H*_0_ values revealing a RMS error below 120% of the minimum were used to estimate uncertainty ranges. These close-to-optimal *H*_0_ were combined with a range in external input *γ*′ varied from 0 to 3 10^3^ (thus two times the reference value, see Tab. S2) to calculate the corresponding uncertainty in model trajectories. Reference settings for *f*_*C*_ >0 were applied in all subsequent experiments apart of a single run without decline in *H* (*r*_*H*_ =0 in Eq.(4), thus *H*=*H*_0_).
B. The calibration in (A) was repeated with the the full data set; the RMS error for the late Dec (2020) to mid Jan (2021) data was weighed ten times higher than for the preceding period in order to achieve a reconstruction at elevated accuracy of the second wave before vaccination started. Also, three global settings of the reference run were systematically calibrated for each region: degradation rate *r*_*H*_, external input *γ*′, and degradation date *t*_reset_. Using the extended parametrization, series of 2-year simulations were conducted with different vaccination scheduling and vaccine effect. Vaccination period Δ*T*_vacc_ was set either to 3 or 9 months to encompass the range of announced plans also accounting for a short immunization period. Vaccine effect describes the acceptance ratio and the (uncertain) vaccine efficacy and was here set to either to 0.7 or 1. Vaccines were in particular assumed to prevent transmission despite lacking evidence so far. Their application followed a common protocol prioritizing the elderly: starting from *i* = 7, the relative fraction of age group *i* was reduced by 1*/*Δ*T*_vacc_ per day until being empty; then *i* was counted down to start with the next cohort.
C. A series of 1.5-year simulations was run across the 20 regions in which *H* was systematically increased from the regional reference value. Import rate *γ*′, vaccination rate, and degradation rate *r*_*H*_ were set zero.
D. Model sensitivity for two regions (Louisiana and Belgium) was assessed by varying 12 parameters 50% up and down from their reference value in Tab. S2.

## Supporting information

SOM

## Data Availability

All used observation were downloaded from publically available sources.
The code required to produce all model results is available at
https://github.com/kaiwirtz/CovidSocMod

## Acknowledgments

The author would like to thank Carsten Lemmen for a critical reading, and Jochen Metzger, Maximilian Schäfer, and Detlef Gronenborn for helpful comments. Apple, Google, and YouGov are acknowledged for making aggregated data available.

## Code availability

The code required to produce all model results is available at: https://github.com/kaiwirtz/CovidSocMod

## Supplementary materials

Supplementary Text

Figs. S1 to S11

Tables S1 to S2

