## Supplementary material for "Decline in mitigation readiness facilitated second waves of *SARS-CoV-2*": SOM

### Supporting Online Material
### for
### ”Decline in mitigation readiness facilitated second waves of *SARS-CoV-2*”

Kai Wirtz

**Supplementary Tables S1 to S2**

Table S1: Regional characteristics: mean latitude ($^o$N), annual minimum and maximum temperature $T_{\min}$ and $T_{\max}$ ($^o$C), resp., median age $A$ (yr) [*1*], relative proportion $h'$ (%) of people above 65yr living in a mixed household [*2*], correction factor of mortality due to excess deaths $f_M$, initial thread awareness $\Delta t$ (d), base human value $H_0$ ($10^4$), and relative immunity $\tilde{S}=1-S$ (%) by May and Dec 2020. Temperature climatology (averaged Feb and Aug temperature 1994-2014) was available for US states [*3*] and non-US countries [*4*]. Regions were ordered with respect to mortality during the first wave.

| country/state | | lat. | $T_{\min}$ | $T_{\max}$ | $A$ | $h'$ | $f_M$ | $\Delta t$ | $H_0$ | $\tilde{S}_{\text{May}}$ | $\tilde{S}_{\text{Dec}}$ |
|---|---|---|---|---|---|---|---|---|---|---|---|
| New Jersey | NJ | 39.8 | 0.0 | 22.6 | 40.0 | 7 | 1.7 | -15 | 1 | 13.9 | 71.0 |
| New York | NY | 43.0 | -4.1 | 20.0 | 39.0 | 7 | 1.1 | -13 | 5 | 9.1 | 64.4 |
| Iran | IR | 32.0 | 6.1 | 31.2 | 31.7 | 30 | $<$15 | -11 | 5 | 3.3 | 25.8 |
| Belgium | BE | 50.5 | 3.8 | 18.3 | 41.6 | 7 | 1.0 | -15 | 7 | 5.8 | 49.8 |
| Massachusetts | MA | 42.4 | -2.7 | 20.4 | 39.5 | 7 | 1.4 | -12 | 2 | 6.6 | 64.9 |
| Italy | IT | 42.5 | 6.6 | 23.4 | 46.5 | 15 | 1.3 | -19 | 10 | 4.4 | 19.6 |
| United Kingdom | UK | 54.0 | 4.4 | 15.1 | 40.6 | 7 | 1.2 | -16 | 12 | 5.5 | 30.5 |
| Switzerland | CH | 47.0 | -0.9 | 14.8 | 42.7 | 7 | 1.2 | -9 | 6 | 2.2 | 28.7 |
| Louisiana | LA | 30.4 | 11.3 | 27.6 | 37.3 | 7 | 1.4 | -7 | 2 | 7.2 | 59.1 |
| Michigan | MI | 44.2 | -5.5 | 19.6 | 39.8 | 7 | 1.2 | -12 | 3 | 4.3 | 53.6 |
| Spain | ES | 40.0 | 7.6 | 23.2 | 43.9 | 21 | 1.2 | -11 | 16 | 5.3 | 27.8 |
| Indiana | IN | 40.3 | -1.0 | 22.6 | 37.8 | 7 | 1.4 | -9 | 4 | 3.0 | 56.7 |
| France | FR | 46.0 | 5.0 | 19.5 | 41.7 | 6 | 1.1 | -11 | 6 | 3.1 | 42.3 |
| Sweden | SE | 62.0 | -7.3 | 13.7 | 41.1 | 2 | 1.0 | -13 | 4 | 4.1 | 29.8 |
| Georgia | GA | 33.2 | 9.4 | 26.3 | 36.8 | 7 | 1.4 | -5 | 4 | 2.6 | 35.8 |
| Netherlands | NL | 52.3 | 3.8 | 17.8 | 42.8 | 3 | 1.2 | -10 | 10 | 3.1 | 25.8 |
| Portugal | PT | 39.3 | 10.2 | 22.7 | 42.6 | 17 | 1.1 | -9 | 14 | 0.9 | 16.4 |
| Germany | DE | 51.0 | 1.6 | 18.2 | 47.8 | 3 | 1.0 | -11 | 13 | 0.5 | 8.9 |
| Ireland | IE | 53.0 | 5.7 | 14.9 | 37.8 | 13 | 1.2 | -15 | 16 | 3.1 | 29.3 |
| Washington | WA | 47.8 | 2.8 | 18.3 | 37.7 | 7 | 1.4 | -12 | 13 | 1.8 | 11.8 |

Table S2: Parameters, auxiliary variables, and dynamic state variables (SV) of the model.

| | Description | Value | Unit |
|---|---|---|---|
| $\alpha_i$ | attack rates | 0.3-1 | |
| $a_y$ | amplitude seasonal fluc. | 0.2 | |
| $A$ | median population age | | yr |
| $\widetilde{A}_i$ | mean age of cohort $i$ | | yr |
| $\beta_0'$ | initial $\beta$ | | $d^{-1}$ |
| $\beta_{ij}$ | age-spec. transmission rates | SV | $d^{-1}$ |
| $\beta_{ij,0}$ | base & initial values of $\beta_{ij}$ | | $d^{-1}$ |
| $C$ | social cost | | $d^{-1}$ |
| $c_H$ | reduction factor of $H$ | 40 | |
| $\delta$ | responsiveness | $1.7\,10^3$ | $d^{-2}$ |
| $\Delta t$ | initial thread awareness | Tab. S1 | d |
| $e$ | effective exposure | | |
| $\epsilon$ | exposure sensitivity | 1.6 | |
| $e_b$ | behavioral exposure | | |
| $e_E$ | environmental exposure | | |
| $f$ | mobility fluctuation | | |
| $\gamma_i$ | external input | | $d^{-1}$ |
| $\gamma'$ | specific external input | $1.5\,10^{-4}$ | |

| | Description | Value | Unit |
|---|---|---|---|
| $h_{ij}$ | household contr. of $\beta_{ij}$ | | |
| $H$ | mitigation readiness | | |
| $H_0$ | base $H$ | Tab. S1 | |
| $I_i$ | infected fraction | SV | |
| $L$ | total loss | | $d^{-1}$ |
| $M$ | COVID-19 mortality rate | | $d^{-1}$ |
| $m_{ij}$ | contact rates | | $d^{-1}$ |
| $\varphi_i$ | age distribution | | |
| $Q_{10}$ | IFR incr. per 10yr age | 3 | |
| $r$ | recovery rate | 0.08 | $d^{-1}$ |
| $r_H$ | degradation rate of $H$ | 0.013 | $d^{-1}$ |
| $r_b$ | learning rate | 0.05 | $d^{-1}$ |
| SD | social distance | | $d^{-1}$ |
| $S_i$ | susceptibles | SV | |
| $t_{\text{reset}}$ | first day of degradation | | d |
| $\theta$ | travel coefficient | 0.35/1 | |
| $\omega_i$ | infection fatality ratio (IFR) | | |
| $\omega_7$ | IFR of oldest group | 0.15 | |

#### S2 Age distribution effects on infection fatality ratio and attack rate

Regional age distributions $\varphi_i$ were interpolated using the median age $A$ from Tab. S1 and the age distributions of the youngest population (Iran, $\varphi_i^0$, with median age $A^0$) and oldest population under consideration (Germany, $\varphi_i^1$, with median age $A^1$):

$$\varphi_i = (1-f)\,\varphi_i^0 + f\,\varphi_i^1 \qquad \text{with} \quad f = \frac{A - A^0}{A^1 - A^0} \tag{S1}$$

Age-specific infection fatality rates (IFR) $\omega_i$ increase in a non-linear way with the medium age $\widetilde{A}_i$ in each cohort, $\widetilde{A}_i \in \{10, 25, 40, 55, 65, 75, 90\}$. Compromising regional studies on the age dependency of case or infection fatality ratios [*5, 6, 7, 8*], I assume an increase by a factor $Q_{10}$=3 every ten years of lifetime:

$$\omega_i = \omega_7 \cdot Q_{10}^{(\widetilde{A}_i - \widetilde{A}_7)/10}$$

where the IFR of the oldest group $\omega_7$ derives from reported estimates ranging from 0.1 to 0.16 [*5, 9, 10, 11*]. Also attack rates $\alpha_i$ are reported to increase with age. Young individuals have a lower $\alpha_i$ than the elderly: they are not only less susceptible, but also less contagious [*12*]. Averaging over results from two studies [*13, 14*] yields the attack rates $\alpha_1$=0.35, $\alpha_{2,3,4}$=0.5 and $\alpha_{5,6,7}$=1.

#### S3 Evaluation of marginal costs and benefits

Marginal costs and benefits of social distancing are in Eq.(7) represented by three pressures that in the model determine responses of societies coping with a deadly virus. The first pressure is generated by the numerous costs caused by social distancing, and mathematically reads as the partial derivative of social cost $C$ with respect to transmission rates $\beta_{ji}$. Using Eqs.(2)–(4) we obtain a term describing relaxation to the pre-pandemic societal life ($m_{ij,0}$):

$$\frac{\partial C}{\partial \beta_{ji}} = 2\,H^{-1} \cdot \frac{\varphi_i\,\varphi_j}{\beta_{ij,0}} \cdot \Big(m_{ij,0} - m_{ij}(\beta_{ij})\Big)$$

The second pressure describes the risk of individuals to become infected and to die depending on their contacts. From Eq.(3) we have:

$$\frac{\partial M}{\partial \beta_{ij}} = \omega_j\,e\,I_i\,S_j \tag{S2}$$

This pressure increases with higher abundance of infected hosts in the interacting group ($I_i$), the size of the (susceptible) target age group ($S_j$), and with its IFR $\omega_j$.

However, the two antagonistic pressures at the level of individual age groups cannot satisfactorily solve the regulation problem for younger age-groups because of vanishing $\omega_j$ and $\frac{\partial M}{\partial \beta_{ij}}$. Therefore, a third term is introduced in Eq.(7) to account for societal pressure as societies seek to lower infection levels of the entire population to better protect risk groups. This motivates the "community-oriented" derivative with respect to aggregated variables in Eq.(7), which contains two terms: First, the marginal dependence of total mortality on total infection density ($\mathrm{d}M/\mathrm{d}I$) equals the IFR. Contrary to the direct partial derivative Eq.(S2), the IFR has to be averaged over all groups: $\sum_k \omega_k \, \varphi_k$. Secondly, the marginal dependence of total infection density $I$ on changes in $\beta_{ji}$ ($\mathrm{d}I/\mathrm{d}\beta_{ji}$) is estimated using a perturbation approach [*15, 16*]. Under steady state conditions we have from Eq.(1)

$$I_i = \frac{\beta_i \, S_i}{r}$$

$$\frac{\mathrm{d}I}{\mathrm{d}\beta_{ji}} = \frac{e \, I_j \, S_i}{r}$$

$$\frac{\mathrm{d}M}{\mathrm{d}I} \cdot \frac{\mathrm{d}I}{\mathrm{d}\beta_{ij}} = \mathrm{e}^{\overline{\beta}\Delta t} \sum_k \omega_k \, \varphi_k \cdot \frac{e \, I_j \, S_i}{r} \tag{S3}$$

The ratio $\beta_i \, S_i / r \, I_i$ is known as reproductive number ("$R$"), which has often guided decision-making during the *SARS-CoV-2* pandemic. Eq.(S3) accounts for a lag period between infection and death, which influenced the estimated case fatalities during the exponential spread phase [*17*]. The mortality rate $M$ in Eq.(3) expresses the actual death toll after a latency, symptomatic, and hospitalization period of approximately 21 days [*17*]. At any time, societies hence perceive a daily mortality which is by the factor $\mathrm{e}^{-\overline{\beta} \cdot 21d}$ smaller than the statistically projected one ($\overline{\beta}$ is the temporal average of recent mean transmission rates).The lag time difference also quantifies the capability to exponentially extrapolate from current case numbers to future mortality rates, which motivates the definition of the "thread awareness" $\Delta t$ in Eq.(S3). $\Delta t$ is in this study only used in the initial part of the simulation as calibration parameter to achieve a synchronous onset of lockdowns across regions (see "Numerical experiments" in Methods). A negative $\Delta t$ corresponds to retarded and indecisive responses by decreasing the community pressure $\frac{\mathrm{d}M}{\mathrm{d}\beta_{ij}}$ responsible for risk reduction.

#### S4 Seasonal mobility cycle

Baseline contact rates ($m_{ij,0}$, Eq.(4), Eq.(S3)) are here assumed to vary with two seasonal factors. First, these changes can reflect the proportion of outdoor vs. indoor activities dependent on thermal comfort as quantified by the temperature function $f_T$:

$$f_T = \mathrm{e}^{-x^2/2} - \frac{1}{2} \qquad \text{with} \quad x = \frac{T - 20^o\mathrm{C}}{5^o\mathrm{C}} \tag{S4}$$

At neutral temperature around 20$^o$C, maximized outdoor contacts will increase $f_T$ ($f_T$=1/2) while at very low or high temperature ($T \ll 10^o$C or $T \gg 30^o$C, respectively), this index will be negative ($f_T$=-1/2). Air temperature $T$ for simplicity follows a trigonometric function including a time lag since in the Northern Hemisphere $T$ is usually lowest around early February:

$$T = T_{\min} + (T_{\max} - T_{\min}) \cdot \sin^2\left(\frac{(t - 35\mathrm{d})\,\pi}{365\mathrm{d}}\right) \tag{S5}$$

with simulation time $t$ counted from January, 1st. Minimal and maximal annual temperatures $T_{\min}$ and $T_{\max}$, respectively, are given in Tab. S1.

As complementary seasonal driver the model includes the annual light cycle quantified by relative daylength DL. DL is calculated using an astronomical formula with time $t$ and the mean latitude of a region (Tab. S1) as inputs. Similar to $f_T$, the seasonal light index $f_{\mathrm{DL}}$ describes how longer days permit more activities and contacts:

$$f_{\mathrm{DL}} = \mathrm{atan}\Big(2 \cdot (\mathrm{DL} - 0.5)\Big) \tag{S6}$$

where the *atan* function smoothes a step function: for locations at higher latitude such as Sweden $f_{\mathrm{DL}}$ approaches -1/2 when DL is small during winter and 1/2 at large DL during summer (most people do not stay outside about 20h per day at summer solstice).

The temperature based index $f_T$ and the light index $f_{\mathrm{DL}}$ are simply added up to yield the overall seasonal mobility fluctuation $f$:

$$f = 1 + a_y \cdot (f_T + \theta f_{\mathrm{DL}}) \tag{S7}$$

The amplitude $a_y$ is estimated from reported seasonal differences in outdoor activities [*18*] and from aggregated mobile phone data for 2016 [*19*]. However, the latter data-set reveals much greater seasonality in Europe compared to the US (SI-Figs.3 and 5 in [*19*]). A much lower seasonal amplitude and maximum intensity in USA compared to European states has also been reported for traveling [*20*]. Therefore, the coefficient $\theta$ in Eq.(S7) is set to one for European states but to a lower value elsewhere (see Tab. S2).

#### S5 Contact vs. transmission rates and reciprocity condition

When considering age-specific mixing patterns, one has to distinguish between transmission rates ($\beta_{ij}$) and contact rates ($m_{ij}$). As introduced in the main text, $\beta_{ij}$ describe the encounter probability per individual, whereas $m_{ij}$ the one per age group. The reciprocity condition ($m_{ij} = m_{ji}$) with Eq.(2) transforms to

$$\varphi_i \beta_{ij} = \varphi_j \beta_{ji}$$

Hence, from the $7{\times}7$ matrix coefficients $\beta_{ij}$, only 28 are independent and used as model variables. To ensure reciprocity at any time, all terms in the evolution equation Eq.(7) are calculated with swapped indices, thus for both $\beta_{ij}$ and $\beta_{ji}$(=$\beta_{ij}\varphi_i/\varphi_j$) and then averaged.

#### S6 Initial and base transmission rates

Business-as-usual (BAU) transmission rates $\beta_{ij,0}$ were retrieved starting from reported (time independent) contact matrices $\overline{m}_{ij,0}$, which were available for European countries from the POLYMOD project [*21*]. In that study, relative proportions of different interaction fields were: 0.3 for households, 0.18 for schools, 0.19 for workplaces and 0.33 for the general community. Analysis of accompanying studies [*21, 22*] suggests that this approach underestimates interaction with friends. I therefore added a 30% or 12% contribution in intra-group contacts for below 30yr or above 30yr, respectively [*23*]. The resulting contact matrices emphasize the diagonal elements (Fig. S11). Contact matrices of non-European countries (Iran, USA) were set equal to the one of UK and transformed to base contact rates $\overline{m}_{ij,0}$ using the interpolated age distribution ($\varphi_i$) described in Sec. S2. Time habits in BAU mixing were considered by multiplication with the seasonal forcing function $f$ given in Eq.(S7):

$$m_{ij,0} = f\,\overline{m}_{ij,0}$$

The date when the reported daily COVID-19 associated death toll meets the threshold $M$=7$\cdot$10$^{-7}$d$^{-1}$ defines the simulation start for each region (see Methods). Then, the initial transmission $\beta_{ij,0}$ (Eq.(2))

$$\beta_{ij,0} = \beta_0' \, \frac{\alpha_i \alpha_j \, f \, \overline{m}_{ij,0}}{\varphi_i}$$

is tuned such that the temporal increase in mortality (proportional to the $\beta_{ij,0}$) during ten days after this onset corresponds to the reported mortality increase. Thus, the proportionality factor $\beta_0'$ is specific for each region (Tab. S1).

#### S7 Minimal contacts depending on household structure

BAU transmission rates $\beta_{ij,0}$ not only define upper target values for the adaptive dynamics in $\beta_{ij}$ in Eq.(S3), they also constitute reference values for lower bounds of social distancing. These lower bounds are especially relevant in simulations at high $H$ since real subsistence activities and household contacts cannot be shut-down entirely:

$$\beta_{ij} \geq \max\{h_{ij}, h_{\min}/h'\} \cdot \beta_{ij,0}$$

Relative household contributions $h_{ij}$ to base transmission rates $\beta_{ij,0}$ not only reflect the global relative weighing factor (0.3, see above) but also the variable age-specific intensity of household interactions. The coefficients $h_{ij}$ were reconstructed from [*24*], with age-dependent co-habitation values of $h_{11}$=0.35, $h_{ii}$=0.45 for $i$=2. . .5, and $h_{66}$=$h_{77}$=0.65, whereas a higher value of $h_{13}$=0.75 is estimated for the intergenerational contact between children and their parents. Apart of children-parents constellations, intergenerational households also comprise the elderly. The relative proportion $h'$ of elderly above 65yr that live in a mixed household has been reported to vary between European countries from 1.6% (SE) to 20.6% (ES) [*2*] (Tab. S1). $h'$ contributes to few non-diagonal coefficients $h_{ij}$ of the household contact distribution: $h_{16}$=$h_{36}$=$h_{37}$=$h_{47}$=$h'$. With a higher fraction of elderly in mixed household I assume a lower relevance of caring infrastructure and also more small-scale solutions for daily supply. Therefore, the subsistence related proportion of contact rates in Eq.(S7) is a constant ($h_{\min}$) divided by $h'$. For example, food supply, nurseries, and health care facilities can hardly be (en)closed in the large absence of care services being provided by families.

In the calculation of the mobility index,

$$\text{Mobility} = \sum_{ij} \Big( m_{ij} - h_{ij} m_{ij,0}/2 \Big) \cdot \varphi_i \varphi_j \tag{S8}$$

for simplicity half of household contacts $h_{ij} m_{ij,0}$ are subtracted from contact rates $m_{ij}$ since many of these contacts will not require routing requests by mobile phone users. Analogous to the mobility index from mobile phone data provided by Google and Apple, the weighed sum was normalized by the weekly average directly after simulation start.

#### S8 Spurious and imported cases

Within each age class, infection density $I_i$ is set to zero when $I_i$ falls below the threshold of $10^{-7}$, which corresponds to one individual in a population of 10 million (to avoid unrealistic residual infection levels). This condition was only met in few scenario runs with very low SCV or towards the end of two year simulations including vaccination.

At low infection density, the (small) external input rate $\gamma_i$ in Eq.(1) gains quantitative importance [25]. The term describes local re-intrusion of cases by business or touristic travelers:

$$\gamma_i = \gamma' \, \varphi_i \, \overline{\beta}_i \tag{S9}$$

with the travel coefficient $theta$ (Sec. S4), a specific global input rate $\gamma'$, the relative size of the age group $\varphi_i$, and contact propensity quantified by the averaged transmission rate $\overline{\beta}_i$=$\sum_j \beta_{ji}/7$. Lacking regional differences, $\gamma_i$ is set to zero before the first lockdown to avoid artifacts in the initial model dynamics.

#### S9 Environmental exposure

The variable exposure $e$ in Eq.(1) describes the effective reduction in transmission independently from social distancing and is in this approach split-up into the two factors behavioral exposure ($e_b$, see Eq.(8)) and environmental conditions ($e_E$)

$$e = e_b \cdot e_E$$

Environmental exposure $e_E$ also carries behavioral aspects but triggered by ambient conditions. Due to the pre-dominant aerosol transmission of *SARS-CoV-2*, outdoor contacts are considered much less contagious so that the effect of temperature driven increase in activity (see above) can be reverted if contacts are shifted from indoor to outdoor micro-environments. These opposing effects may also explain lacking clear temperature correlations. However, many studies point to an at least moderate decrease in transmission efficiency with rising temperature [*26, 27, 28, 29, 30*]. Underlying these findings may be decreased viability of *SARS-CoV-2* on surfaces at higher temperature [*31, 32*]. More pronounced are effects of UV light on viral activity [*33, 34*]. In short, summer conditions ($f > 1$, see Sec. S4) have a strong negative impact on transmission efficiency, both indirectly and directly. As a consequence of direct ambient forcings, seasonal fluctuations not only revert their sign compared to the formulation $f$ in Eq.(S7), but also increase in magnitude:

$$e_E = 1 - 2a_y \cdot (f_T + \theta f_{\mathrm{DL}})$$

The increase of activities and contacts in warmer and lighter seasons is thus more than compensated by a higher reduction in environmental exposure $e_E$. This setting and parametrization was already condensed in a preceding model study to a net effect of a maximal temperature variation on transmission of around 25% [*35*].

#### S10 Model limitations and uncertainties

Uncertainties of this reconstruction first pertain integrated data. The mortality data only roughly account for excess deaths or for region-specific or shifting attribution criteria such as for Ireland on Apr, 24. Although trends in the mobility data from Apple could be confirmed by similar products provided by Google (not shown) or national providers [*36*], the association between routing requests and real physical contacts remains approximate. Yet, aggregate mobility seems to be a better proxy than implementation dates of certain policies since their containment effect can differ between countries [*37*]. Future model versions should include an updated parametrization and more details from existing SIR models such as spatial structure [*38, 39, 40*] or age dependency of infectiousness [*41*]. The aggregate nature of the societal model should be dis-entangled to better resolve partially independent dynamics in political, economical, cultural, and psychological constituents of behavioral exposure and mitigation readiness. Aggregation also infers a lower spatial limit of model applications as in local communities more specific drivers can be expected to drive mitigation dynamics. The aggregation finally imposes a lower temporal scale as it neglects, among others regional differences in cultural conditioning. For example, in Ireland or Georgia, mitigation readiness likely dropped during Christmas, leading to massive mortality peaks, which were predicted by the model in terms of magnitude but with a relatively large time delay.

A sensitivity analysis demonstrated qualitative robustness of major outcomes (Fig. S10), but also revealed few major uncertainties such as for IFR or attack rates of the younger age classes, or for the few societal coefficients of the model. Certain model settings and assumptions will probably remain untested since changing behavior during crises situation is difficult to monitor. Due to the lower median age of the US compared to European countries (Tab. S1), the here presented selection of regions infers a weak correlation between median age and $H$, which may not be the case for emerging countries (e.g., see intermediate $H$ of Iran).

#### Supporting figures S1–S11

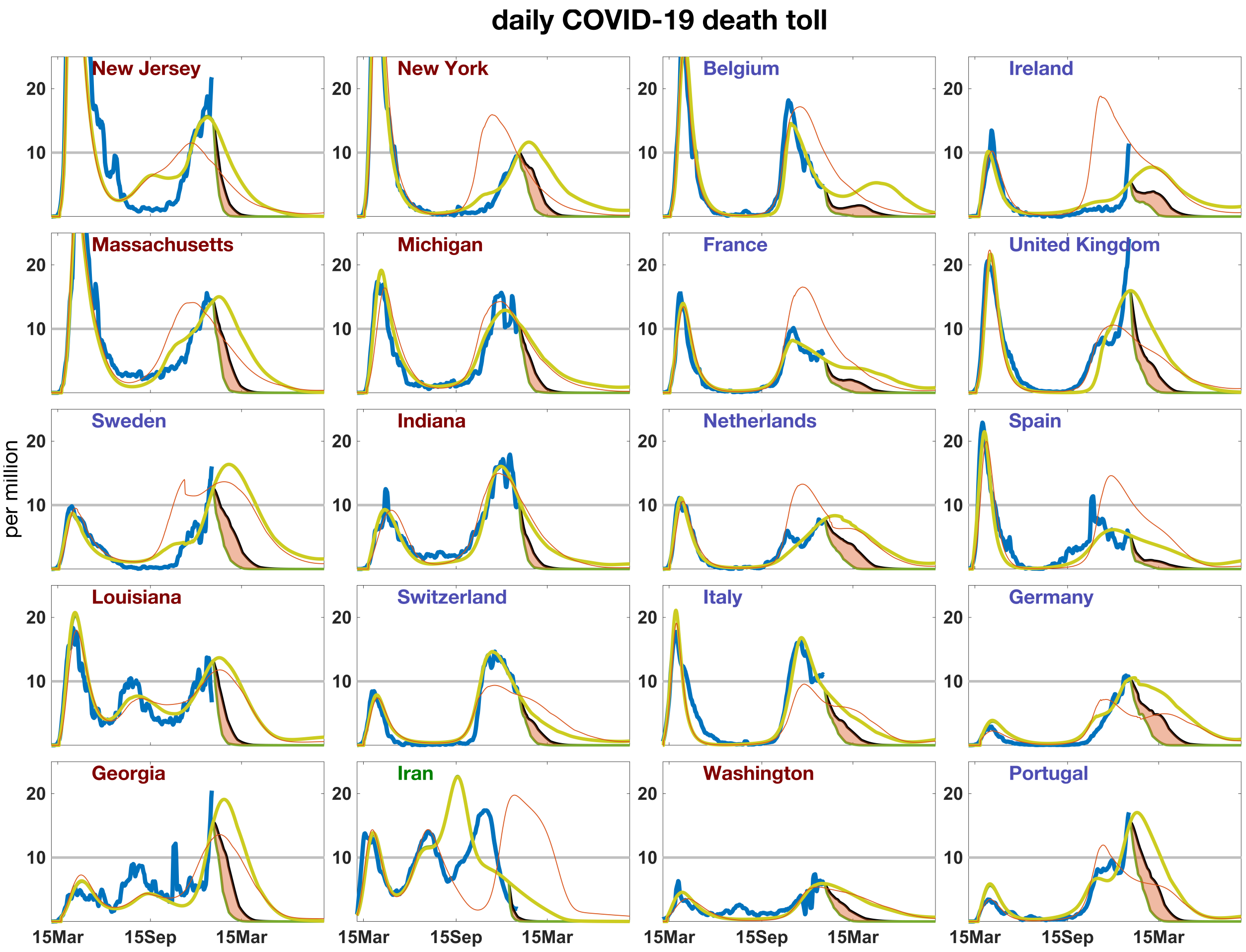


Figure S1: Fig. 2 Mortality rate in the reference simulation (thin red line) compared with reported data (blue line) and a simulation with extended calibration (olive line) where three parameters were fitted for each region separately using the full data set instead of only the first 180d. The mortality difference between a 3-months vaccination scheme (green line) and a 9-months scheme (black line), both at 100% efficacy, is visualized as red shading. Calibrated values for degradation rate $r_H$ ($10^{-2}$ d$^{-1}$, Eq.(5)), external input $\gamma'$ ($10^{-4}$, Eq.(1) and S9), and degradation date $t_{\rm reset}$ (day in 2020, Eq.(5)) are: BE:1.1/0/321, FR:0.5/2/193, DE:4.4/2/289, IR:3.3/0/193, IE:3.3/12/289, IT:4.4/2/225, NL:2.2/5/257, PT:3.3/2/289, ES:1.7/12/161, SE:3.9/5/289, CH:3.3/2/225, UK:3.9/0/289, GA:2.8/0/289, IN:2.8/12/225, LA:2.2/5/257, MA:2.8/0/289, MI:1.7/10/225, NJ:5.0/0/289, NY:2.8/12/289, WA:4.4/12/257.

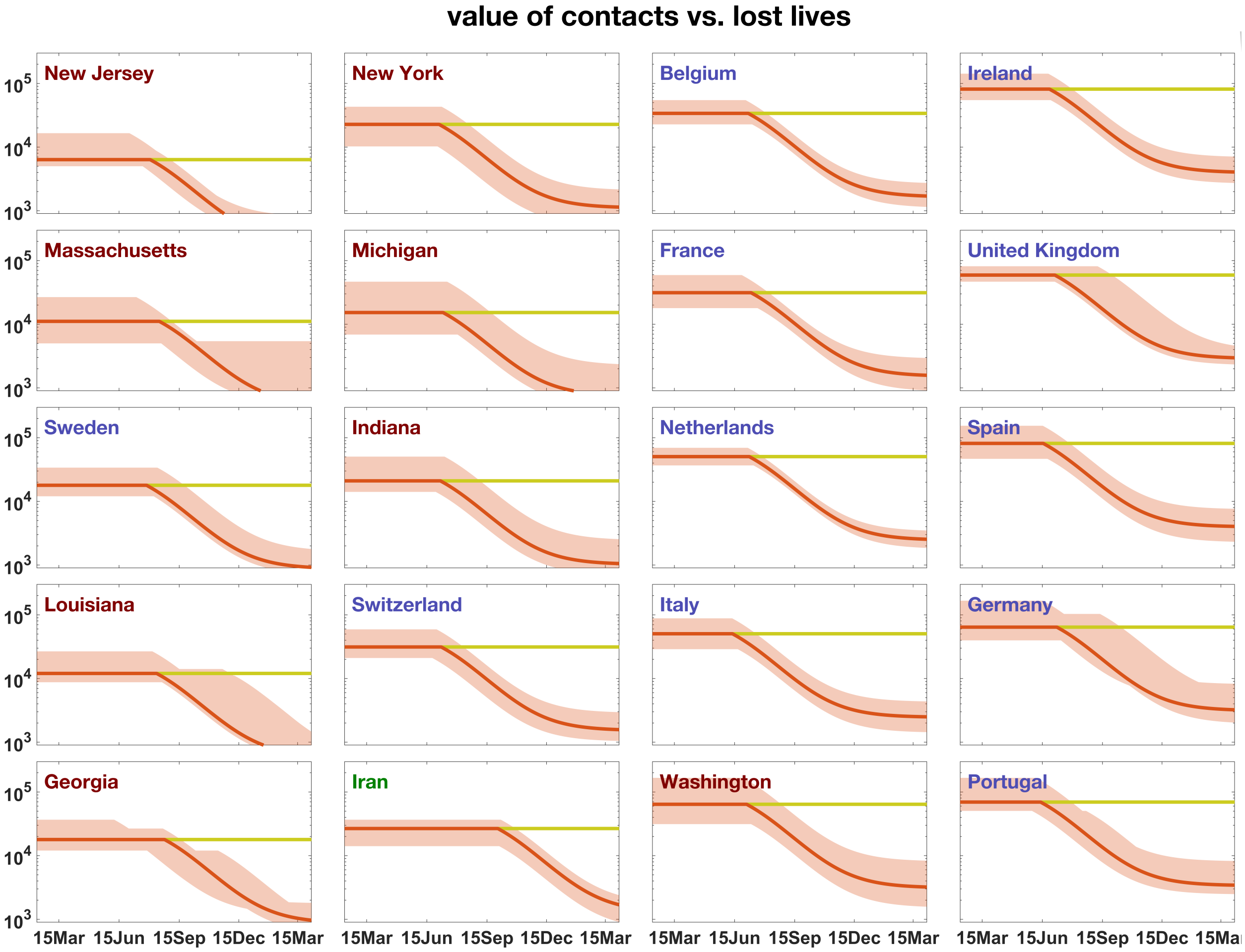


Figure S2: The value of human lives versus BAU contact rates, here denoted as mitigation readiness $H$, in the two model variants: either with degradation of $H$ from its regional base value $H_0$ ($r_H > 0$ in Eq.(5), red line), or without degradation (olive line). Uncertainty ranges in the variant with degradation (shaded area) derive from a released selection criteria during parameter calibration (see Methods).

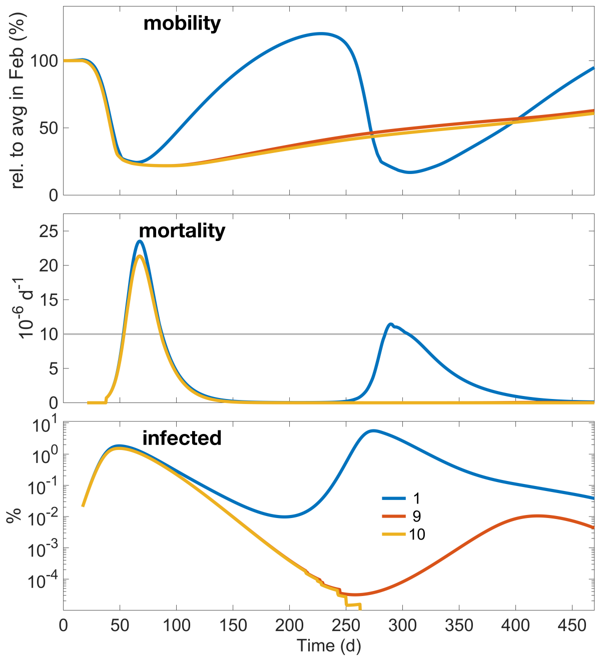


Figure S3: Comparison of aggregated variables in simulations for Belgium where the mitigation readiness is kept constant within the run ($H$=$H_0$), but varied for each run from the reference value ($H_0$=6.8 $10^4$). Three proportionality factors were used: 1 (blue line), 9 (red), and 10 (orange). From top: mobility from Eq.(S8), mortality $M$, and total fraction of infected individuals $I = \sum_i I_i$.

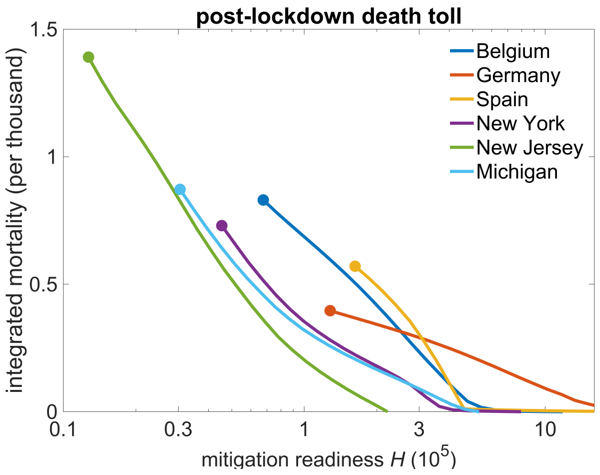


Figure S4: Death toll after the first wave depending on the base mitigation readiness ($H$=$H_0$) for six regions. Reference $H_0$s are highlighted as circle; travel input and degradation were switched off ($\gamma'$=0, $r_H$=0).

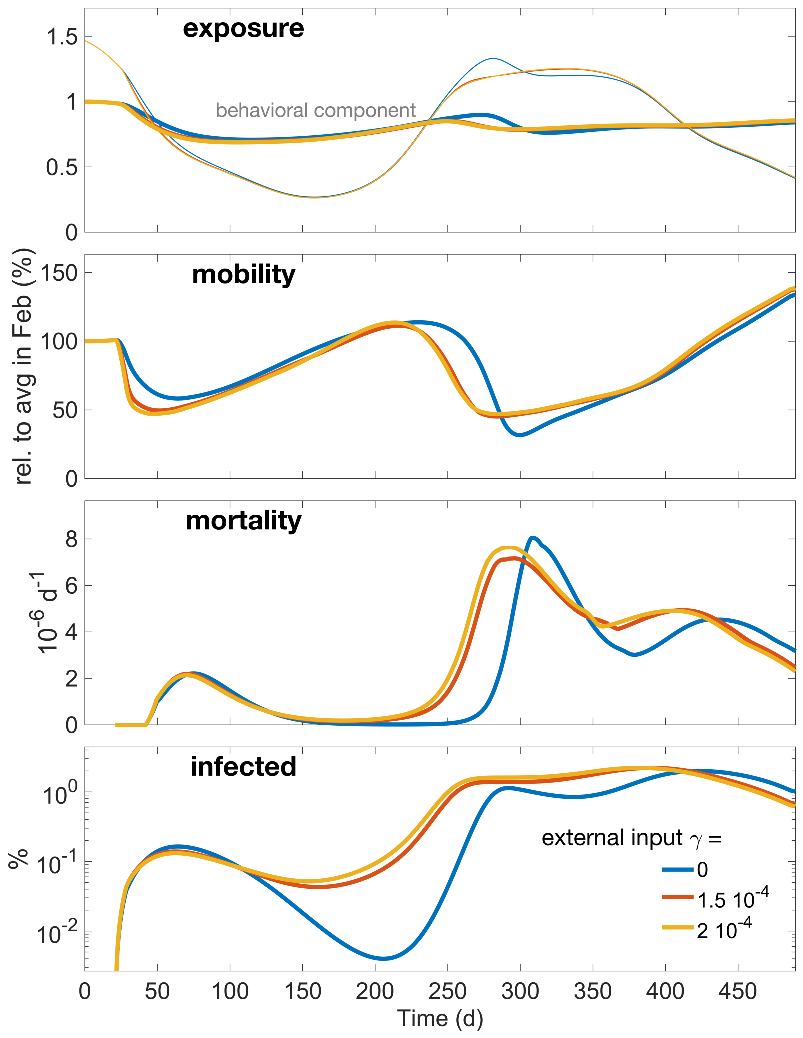


Figure S5: Aggregated variables in simulations where the reference value of the specific external input $\gamma'$=1.5 $10^{-4}$ (red line) is varied either downward ($\gamma'$=0, blue line) or upwards $\gamma'$=2 $10^{-4}$ (orange line). From top: effective exposure $e$ (thin lines) and the behavioral contribution $e_b$ (thick lines), mobility, mortality, and relative number of cases in the population.

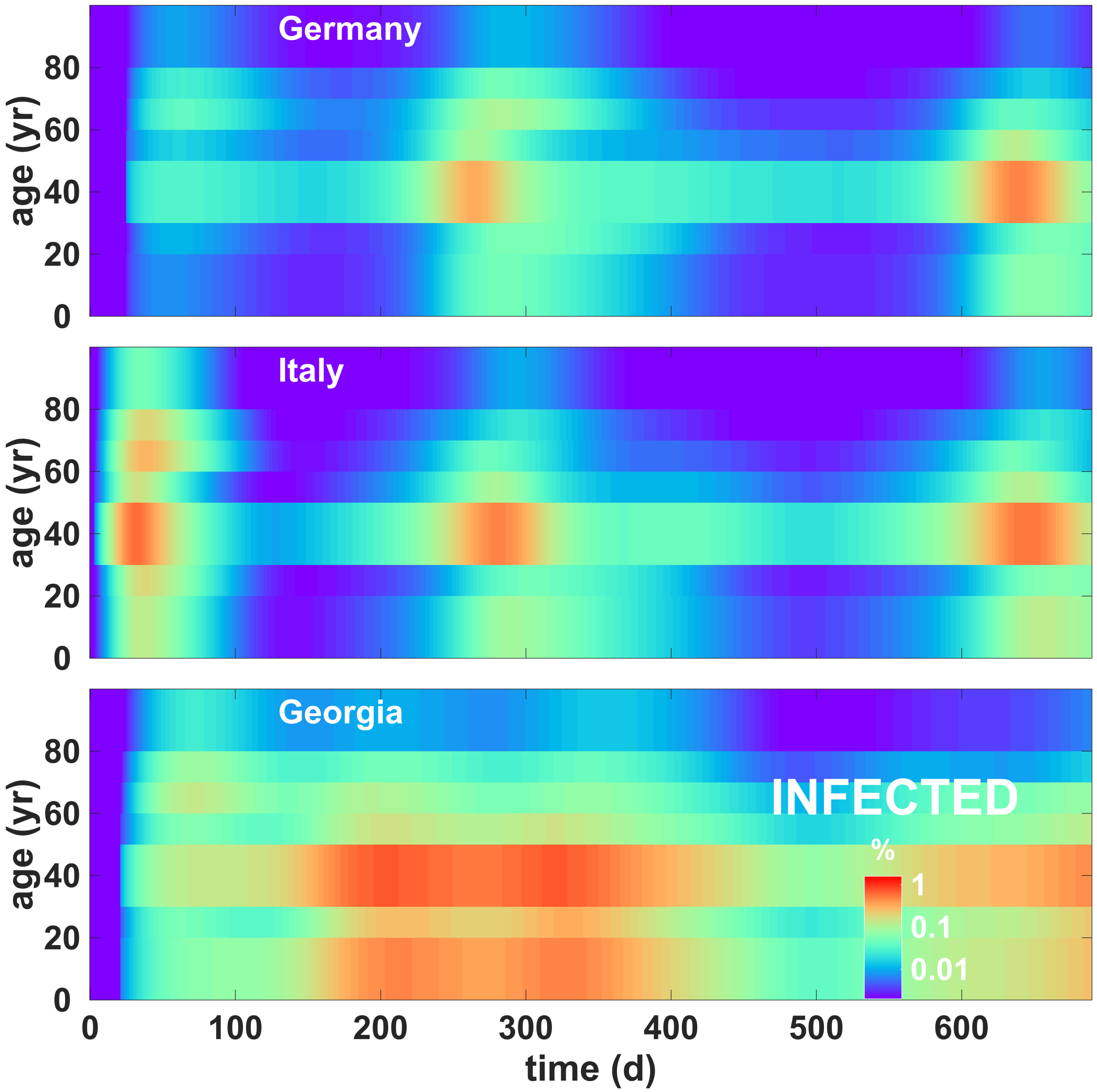


Figure S6: Time evolution of the relative density of infected people ($I_i$) over two years for all age classes in Italy, Germany, and Georgia in case of no vaccination.

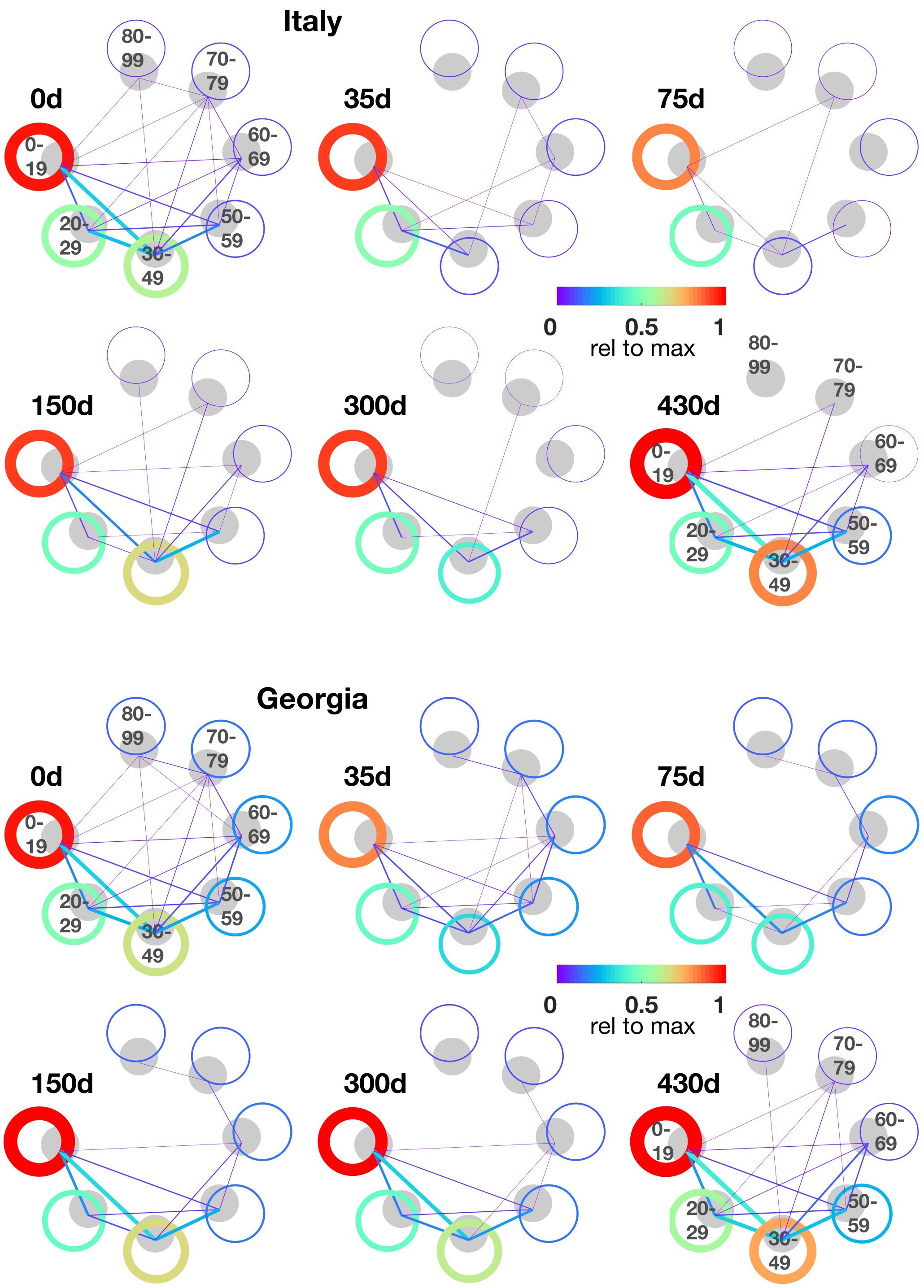


Figure S7: Age-specific interaction patterns ($\beta_{ij} \cdot \varphi_i$) at distinct times in Italy and Georgia. Contact intensity is normalized to the pre-pandemic interaction in the youngest age group and represented as color and line thickness.

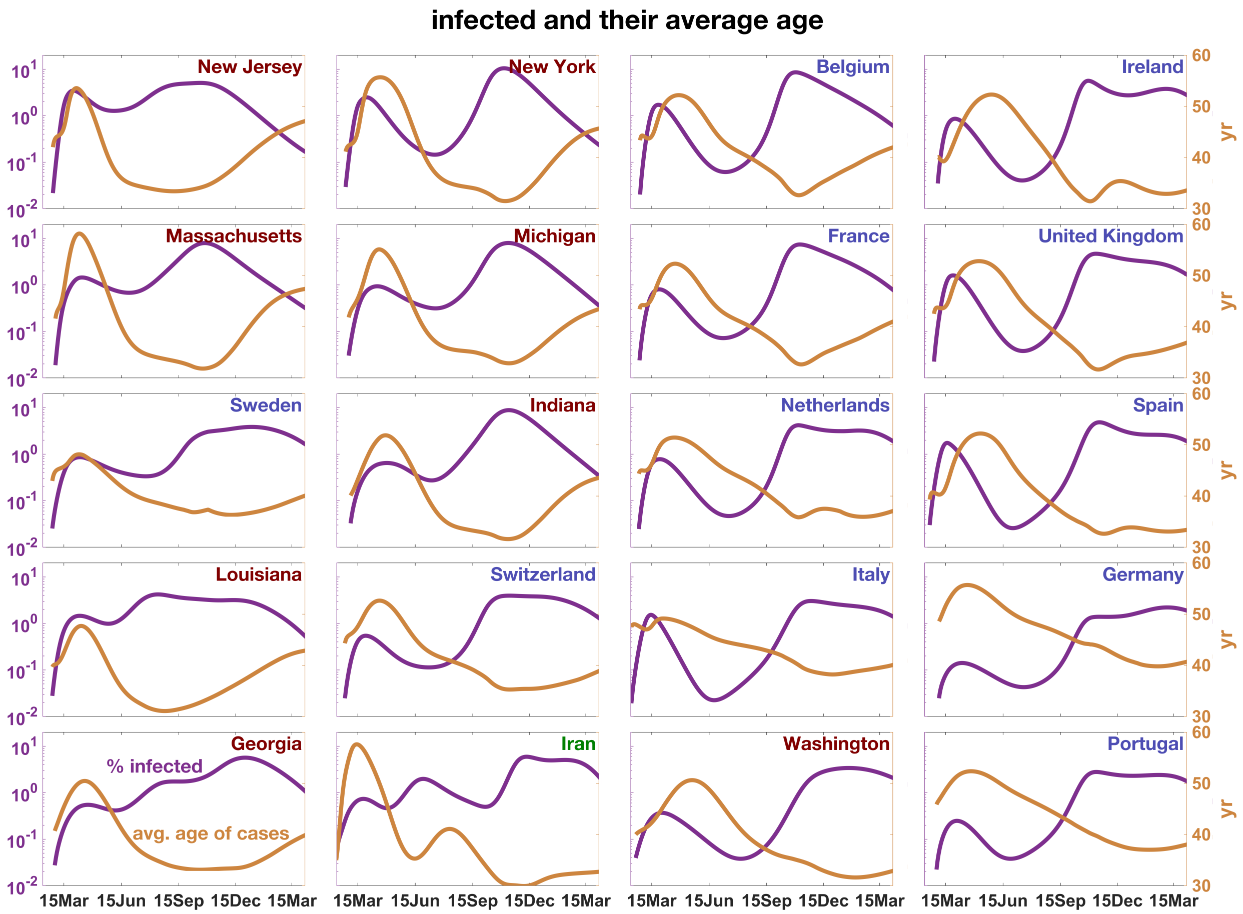


Figure S8: Simulated fraction of infected cases $I = \sum_i I_i$ (purple line) and their mean age (brown line) for the 20 study regions from Feb 2020 to Apr 2021 (reference run with $r_H > 0$ and without vaccination).

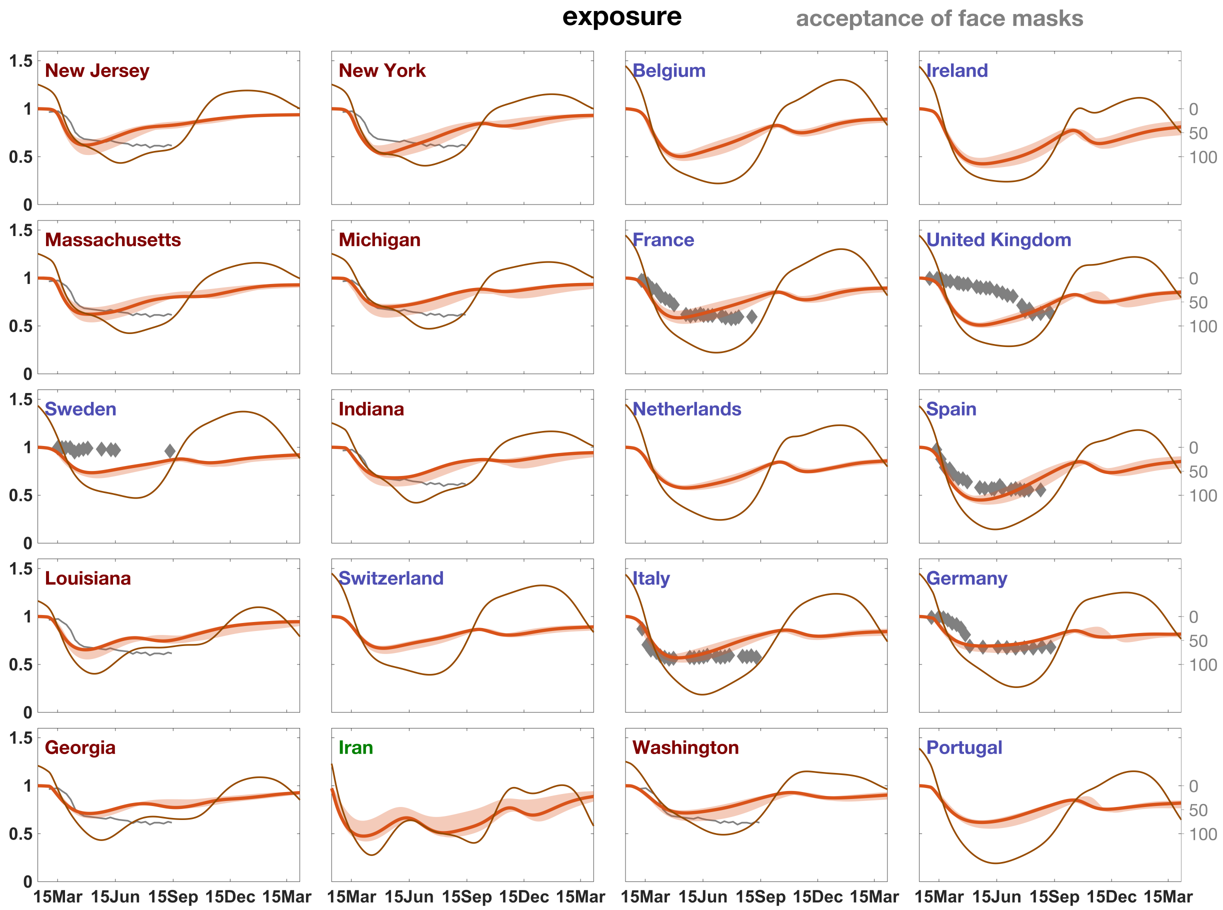


Figure S9: Behavioral exposure $e_b$ (red line) and effective exposure, $e = e_b \cdot e_E$ (brown line), thus encompassing the seasonality of the environmental exposure $e_E$, simulated for all study regions. Behavioral exposure is tentatively compared with the survey data on the acceptance to wear face masks in the public [*42*] (grey dots; willingness in %). For the US, no data were available at state level. European countries are labelled in blue, US states in red.

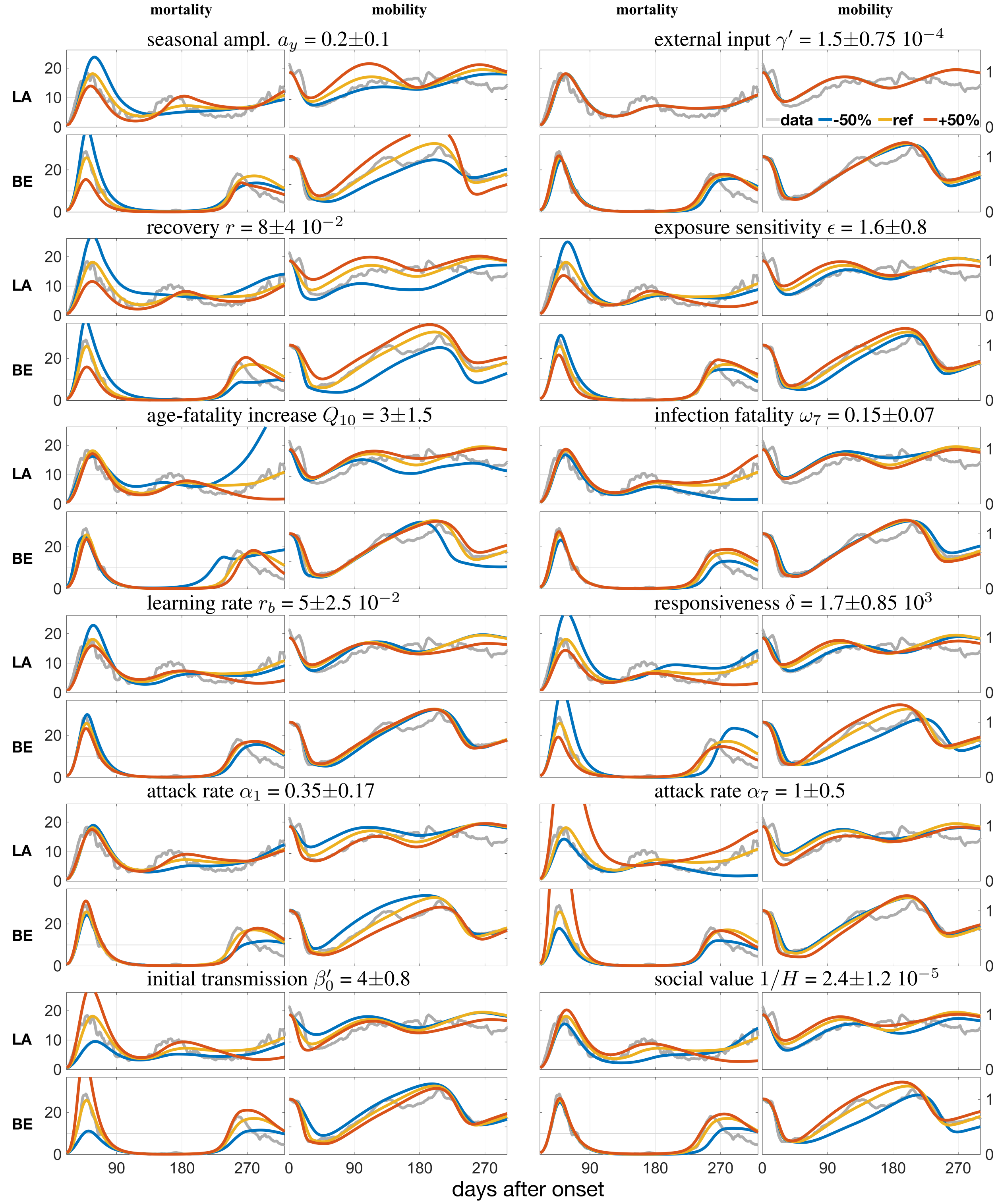


Figure S10: Sensitivity analysis: effect of individual parameter variations on two major model variables (mobility and mortality) for Belgium (BE) and Louisiana (LA).

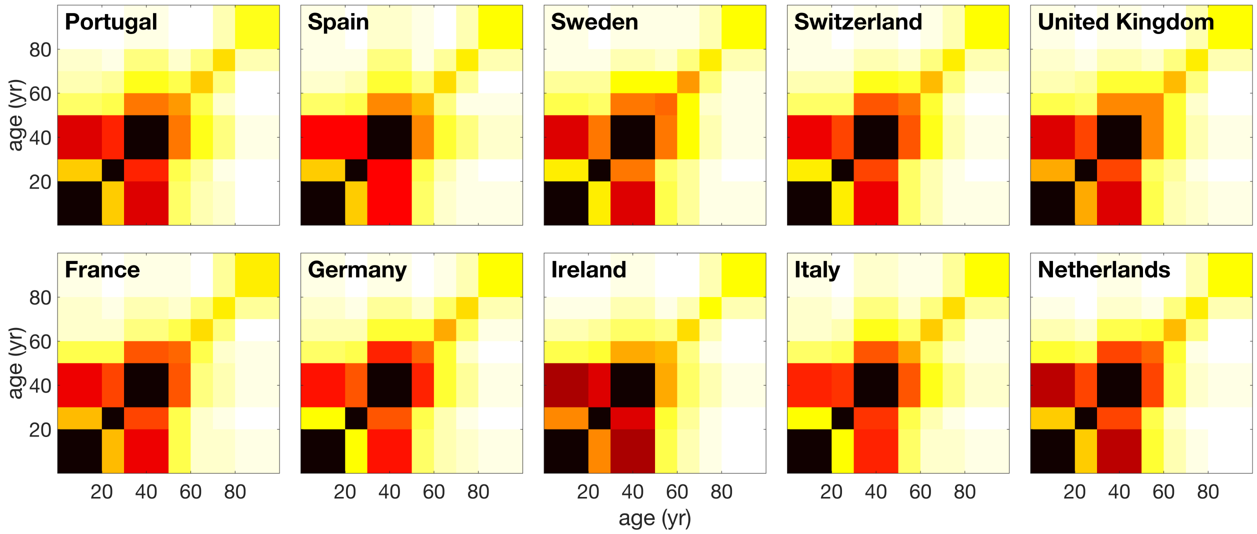


Figure S11: Initial and BAU contact matrices $m_{ij}$ for ten European countries from POLYMOD data [*21*] after adding intra-group contacts with friends. Dark fields indicate strong mixing.
